# Biological drifts within normal ranges allow the detection of Crohn’s disease patients at high risk of rehospitalization

**DOI:** 10.64898/2026.07.21.26358574

**Authors:** Alexandre Homo, Jakez Rolland, Clément Bézier, Ronan Boutin, Leila Equinet, Nathalie Maes, Marie Thys, Lucie Monin, Edouard Louis

## Abstract

**Background:** Crohn’s disease is a chronic relapsing inflammatory bowel disease with an unpredictable clinical course that may lead to recurrent hospitalizations and surgery, making early identification of patients at risk a key challenge in longitudinal monitoring.

**Objective:** To evaluate the prognostic value of blood biomarkers for anticipating hospitalizations in patients with Crohn’s disease by moving beyond exclusive reliance on conventional reference intervals toward the analysis of personalized biological drift. The underlying premise is that fluctuations that remain within standard reference ranges (and are therefore invisible to conventional thresholds) may still carry a risk signal when interpreted relative to an individual’s optimal baseline.

**Design:** We conducted a retrospective study of 993 patients with Crohn’s disease followed at the University Hospital of Liège between 2005 and 2023. Longitudinal laboratory measurements were linked to Crohn’s disease-related hospitalizations. Biomarkers were transformed into z-scores relative to optimized and personalized reference populations and classified into drift categories. Time to first hospitalization was analyzed using the Kaplan-Meier method, and recurrent hospitalizations were modeled using Cox models.

**Results:** Hospitalization-free survival differed significantly across drift categories, including for deviations within conventional reference ranges (e.g., albumin, global log-rank p<0.0001). Among 57 biomarkers screened, 32 were significant in the global log-rank analysis, including 5 that were significant for intra-reference drift classes: low lymphocytes (%), low monocytes (%), low albumin, high potassium, and low aspartate aminotransferase.

**Conclusion:** Personalized biomarker drift detects clinically meaningful risk signals that are missed by conventional reference-interval thresholds and may enable earlier risk stratification in Crohn’s disease.

**Key messages:** *What is already known about this topic:* Routine monitoring of Crohn’s disease relies on inflammatory markers and conventional reference intervals, but relapse prediction remains difficult, and subtle within-range changes in common blood biomarkers remain poorly characterized.

*What this study adds:* In a real-world longitudinal cohort of 993 patients, personalized biological drift categories identified hospitalization risk signals in routine biomarkers, including albumin, lymphocytes, monocytes, potassium, and aspartate aminotransferase, that would be missed by standard thresholds.

*How this study might affect research, practice, or policy:* These findings support further evaluation of personalized reference-population approaches for routine blood tests, with the aim of improving early risk stratification and dynamic monitoring in Crohn’s disease.

## Introduction

Inflammatory bowel diseases are highly prevalent worldwide and their incidence has been steadily increasing throughout the 21st century (1). Ulcerative colitis and Crohn’s disease are the most common forms, together affecting an estimated 6 to 8 million people globally in 2017 (2). These conditions impose significant medical and economic burdens, which are often difficult to quantify at a global level (3).

Crohn’s disease (4) is a chronic relapsing inflammatory disorder of the gastrointestinal tract, driven by dysregulated immune responses and associated with progressive mucosal damage, including ulceration (5). Its clinical course is marked by alternating periods of remission and flare-ups with highly variable frequency and duration (6), making longitudinal management complex and requiring continuous adaptation of treatment strategies. Although many patients can be managed in ambulatory care, acute exacerbations, complications, or treatment failure frequently lead to hospitalization and, in some cases, surgery (7, 8). In this context, therapeutic escalation may involve immunosuppressants, targeted biologics, and surgical interventions when irreversible bowel damage occurs (9). Therefore, anticipating relapse and preventing severe outcomes remain central goals of Crohn’s disease monitoring.

As part of follow-up in diagnosed patients, laboratory tests are prescribed to monitor specific biomarkers such as fecal calprotectin (FC), C-reactive protein (CRP), and other less specific blood markers, including complete blood counts and liver function tests. While CRP is commonly used as an inflammatory biomarker in this context, its specificity for Crohn’s disease remains limited (10). Similarly, although FC is a relevant tool for assessing intestinal inflammation, its implementation in clinical practice is considerably more demanding than a simple blood test (11). Routine blood biomarkers have not been extensively studied as potential prognostic markers of disease progression, with the exception of the neutrophil/lymphocyte ratio (12). Predicting relapse in patients with Crohn’s disease remains challenging (13), but certain biomarkers may reveal weak signals that could alert clinicians and help guide follow-up (14).

The present study aims to evaluate the relevance of blood biomarkers available in real-world practice, including variation within reference ranges, for rehospitalization risk stratification in Crohn’s disease. This is a retrospective longitudinal study of 993 patients with Crohn’s disease followed between 2005 and 2023 at the University Hospital Center of Liège (UHCL). Biological data were analyzed using an innovative methodology developed by Bio Logbook to optimize and personalize laboratory test interpretation (15). Using reference populations to contextualize patient profiles enables a more refined and individualized interpretation of biological variation beyond conventional reference intervals (RIs).

## Materials and Methods

### Outcome definition

Hospitalizations were defined as the primary outcome and categorized by whether surgical intervention was required and by frequency (none, a single hospitalization, or multiple rehospitalizations). Hospitalizations with surgery included any digestive surgical procedure performed in relation to Crohn’s disease (e.g., intestinal resections, stoma creation, abscess drainage). In contrast, hospitalizations without surgery corresponded to admissions for inflammatory flare-ups, medical complications, or therapeutic adjustments without associated operative procedures but still directly linked to Crohn’s disease.

### Statistical analysis of biological data

For each biomarker in patients with Crohn’s disease, values were transformed into z-scores by comparison with optimized and personalized reference values derived from retrospective data collected at a community laboratory in Montpellier, France (15) (see Supplementary Notes for the cohort description), and recalibrated using the Liège University Hospital population.

Reference populations were selected using the following procedure. Let *Y* represent a biological parameter within the selected reference population and *X* denote another biological parameter or a subject-specific characteristic (e.g., sex, age). For each *Y*, the correlated *X*_*i*_ variables were identified using stepwise ANOVA. Among a cohort of *N* individuals with *k* covariates *X* (including biological parameters and/or subject-specific characteristics), reference individuals were defined as those for whom all *Y*-correlated laboratory parameters fell within their respective 95% RIs. These selected individuals were then further stratified based on subject-specific characteristics. Ultimately, a subset of *N*_0_ (≤ *N*) reference individuals was established for each subgroup. We refer to such a subset as a personalized and optimized reference population because it comprises individuals who are more homogeneous and less affected by biological variation than those in the global population. Relevant thresholds were then derived from these populations. The corrected mean, obtained after Box-Cox transformation of the reference values, was considered the “personalized optimum,” while the quantiles {2.5%, 15.8%, 84.1%, 97.5%} and {68.4%, 95%} were used to stratify values for bilateral parameters (with both low and high thresholds) and unilateral parameters (with only a high threshold), respectively (see Table 1). For a given patient, biological values can be compared with the deviation levels of a personalized reference population (with the same subject-specific characteristics) to assess whether they are within expected limits.

**Table 1.** Classification of patients according to biological drift.

| **Biological Drift** | **Bilateral biomarkers**<br>**(percentiles)** | **Unilateral biomarkers**<br>**(percentiles)** |
| --- | --- | --- |
| BRI* | NA | NA |
| Very Low Drift (VLD) | value < 2.5th percentile | NA |
| Low Drift (LD) | 2.5th percentile < value < 15.8th percentile | NA |
| Optimal (Opt) | 15.8th percentile < value < 84.1st percentile | value < 68.4th percentile |
| High Drift (HD) | 84.1st percentile < value < 97.5th percentile | 68.4th percentile < value < 95.0th percentile |
| Very High Drift (VHD) | value > 97.5th percentile | value > 95.0th percentile |
| ARI** | NA | NA |

\*BRI: Below Reference Interval; \*\*ARI: Above Reference Interval

Each individual in the Crohn’s disease cohort was assigned to an optimized and personalized reference population based on sex and age. Biomarker values were standardized relative to the corrected mean of the corresponding reference population, expressed in terms of the number of reference population standard deviations (z-score), and classified into drift classes (Table 1). This approach accounts for between-subject heterogeneity and standardizes the results according to personalized optima and variation.

### Survival modeling

To fully leverage longitudinal laboratory data, each patient could contribute multiple measurements over time and could therefore belong to different drift classes during follow-up. This strategy increases statistical efficiency but introduces within-patient dependence across observations.

We first screened all available biomarkers for association with time to first Crohn’s disease-related hospitalization (first hospitalization event) using Kaplan-Meier analyses (16), initially in univariate models and then in bivariate models adjusted for CRP status (ARI: >5 mg/L vs Opt: ≤ 5 mg/L). A global log-rank test (17) was used to test for global differences across all drift classes for the 57 parameters tested. For significant parameters, we performed pairwise log-rank tests with the Opt class as the reference. All p-values were corrected for multiple testing using the Benjamini-Hochberg method.

Biomarkers with significant drift levels in Kaplan-Meier models and log-rank analyses were then entered into recurrent-event Cox models (18). Rehospitalization recurrence was modeled using Andersen–Gill (AG) (19), Prentice–Williams–Peterson total-time (PWP-TT), and Prentice–Williams–Peterson gap-time (PWP-GT) approaches (20) to account for event-order dependence and time since the previous event. Furthermore, to capture unobserved heterogeneity between patients, we implemented each of these models both with and without a frailty component (21). We focused principally on the PWP-GT model with frailty because it models the time between recurrent events; the other models were used to assess global robustness.

For each biomarker, Cox analyses were performed using two stratification strategies: (i) with drift levels: each drift class (including ARI and BRI) was compared to Opt; (ii) without drift levels: only ARI and BRI were compared to Opt (to simulate current clinical practice based on RIs). In this setting, all values within the reference interval (WRI) were defined as the Opt class.

Finally, we assessed interactions between drift levels and hospitalization type (with vs. without surgery). Additional details on the model rationale and choice criteria are provided in the Supplementary Notes.

### Patient and public involvement

Patients were not involved in the design or conduct of the study, but the results will be published open access to make them available to patients.

To facilitate interpretation of the analytical framework, Figure 1 provides a summary of the research protocol.

**Fig. 1.**
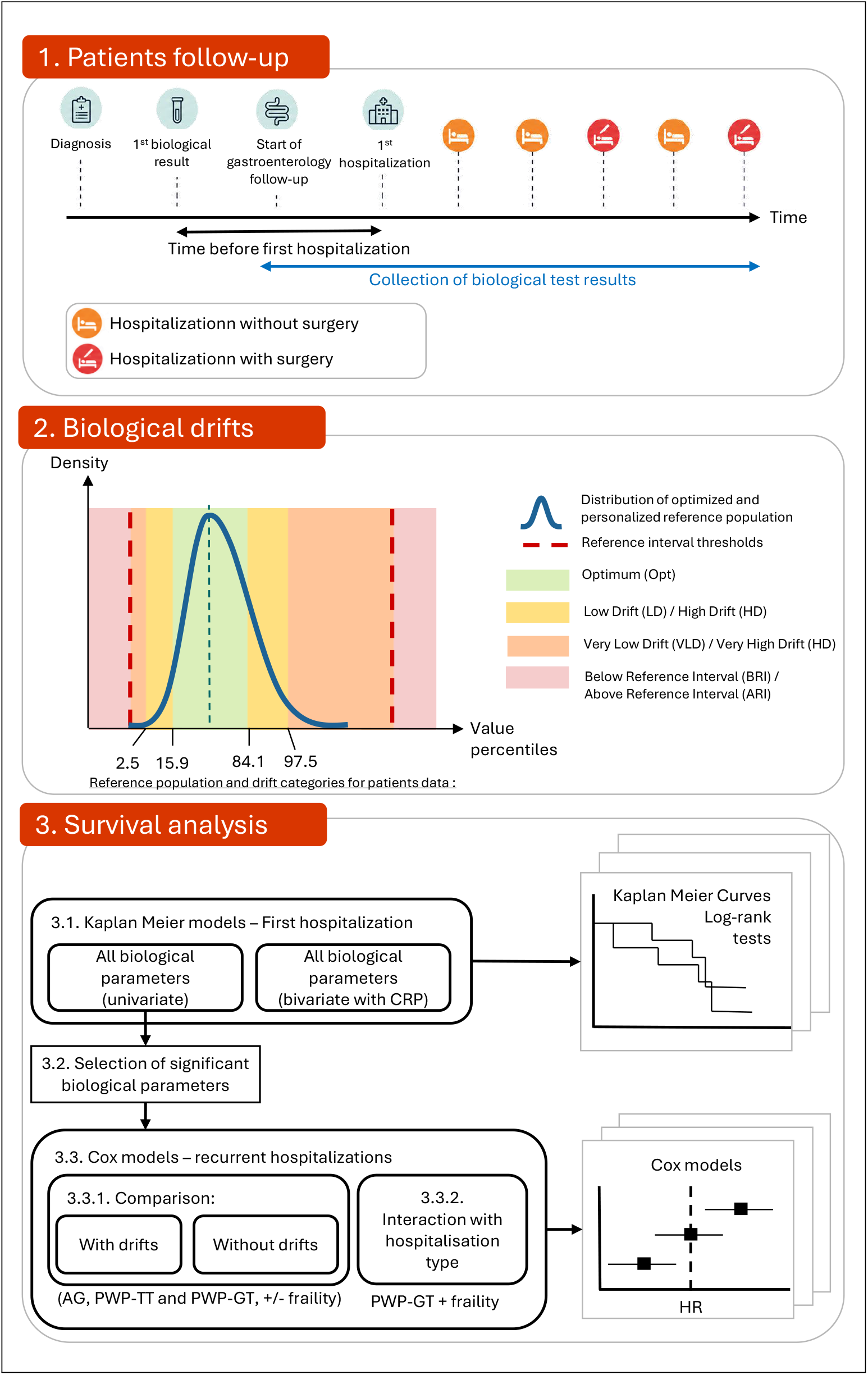
Overview of the research protocol. 1. Patient follow-up: schematic overview of patient inclusion and the analysis period (see Materials and Methods, section *Outcome definition*). 2. Biological drift: schematic distribution of a hypothetical optimized and personalized reference population. For a new result, the biological drift level is determined by comparison with the percentiles of the reference population (see Materials and Methods, section *Statistical analysis of biological data*). Opt: Optimum; VLD: Very Low Drift; LD: Low Drift; HD: High Drift; VHD: Very High Drift; BRI: Below Reference Interval; ARI: Above Reference Interval. 3. Survival analysis: 3.1. Kaplan-Meier models with log-rank tests, 3.2. Selection of significant biomarkers based on corrected p-values, 3.3. Cox models with 3.3.1. Comparison between models with drift-level categories and models with only BRI-WRI-ARI classification (all models tested with or without a frailty component; AG: Andersen-Gill; PWP: Prentice-Williams-Peterson; GT: Gap Time; TT: Total Time), 3.3.2. PWP-GT model with frailty and hospitalization type (with surgery [A] or without surgery [B]) as an interaction term. See Materials and Methods, section *Survival modeling*.

## Results

### Cohort description

A retrospective cohort of 993 patients with Crohn’s disease followed at the UHCL between 2005 and 2023 was assembled. Patients were included if they had a confirmed diagnosis of Crohn’s disease and available longitudinal biomarker records. For each patient, Crohn’s disease-related hospitalization data were linked with biological test results collected throughout follow-up. Hospitalizations were frequent: 58% of patients required at least one hospital admission related to Crohn’s disease, and 35% of patients were hospitalized twice or more during the follow-up period. Surgical intervention was also required in a considerable proportion of cases, with 40.5% of patients undergoing at least one operation over the course of follow-up (see Table 2). The recurrence of hospitalization events is shown in Supplementary Figure 1 (in 30 randomly selected patients from the cohort) and discussed in the Supplementary Notes.

**Table 2.** Cohort characteristics.

| Specifications | Total patients (N=993) |
| --- | --- |
| **Sex** |  |
| Female | 595 (59.9%) |
| Male | 398 (40.1%) |
| **Age at diagnosis (years)** |  |
| 0-20 | 195 (19.6%) |
| 20-40 | 368 (37.1%) |
| 40-60 | 80 (8.1%) |
| 60-80 | 19 (1.9%) |
| 80+ | - |
| No information | 331 (33.3%) |
| **Age at follow-up start (years)** |  |
| 0-20 | 130 (13.1%) |
| 20-40 | 507 (51.1%) |
| 40-60 | 278 (19.6%) |
| 60-80 | 77 (19.6%) |
| 80+ | 1 (0.1%) |
| No information | - |
| **Age at follow-up stop (years)** |  |
| 0-20 | 10 (1.01%) |
| 20-40 | 406 (40.9%) |
| 40-60 | 352 (35.4%) |
| 60-80 | 200 (20.1%) |
| 80+ | 25 (2.52%) |
| No information | - |
| **Number of patients with hospitalization** |  |
| No | 419 (42.2%) |
| Yes | 574 (57.8%) |
| **Number of patients by type of hospitalization** |  |
| Surgery | 193 (33.6%) |
| No surgery | 209 (36.4%) |
| Both | 172 (30%) |
| **Number of patients by number of hospitalizations** |  |
| 0 | 419 (42.2%) |
| 1 | 225 (22.7%) |
| 2 | 137 (13.8%) |
| 3 | 69 (7.0%) |
| 4 or more | 143 (14.3%) |

### Case study: Albumin follow-up

Although they remain strictly within conventional reference thresholds, some biomarkers display notable dynamic variation over time. This is particularly the case for albumin, whose trajectory shows intra-threshold fluctuations when examined in the context of a patient’s longitudinal follow-up. The albumin follow-up for one patient is shown in Figure 2.

**Fig. 2.**
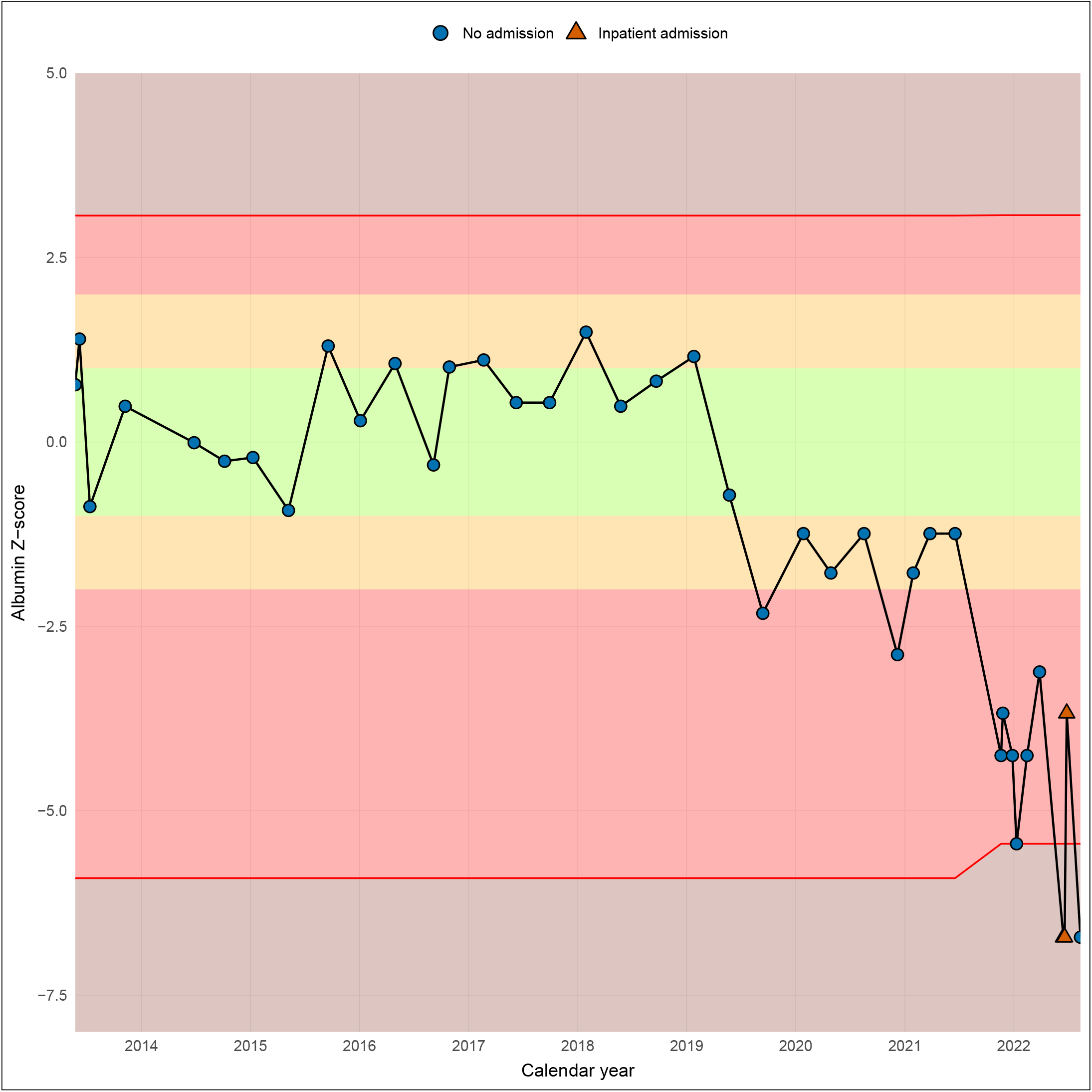
Follow-up of albumin levels in a Crohn’s disease patient. Albumin values have been standardized as z-scores by comparison with the reference population. The green area represents this patient’s personalized optimal range (Opt), the orange area indicates the drift zone (LD and HD), the red area corresponds to the critical zone (VLD and VHD), and the brown areas denote abnormal results according to conventional reference values (BRI and ARI).

These variations are not necessarily considered abnormal according to standard RIs, yet they reflect instability or a loss of fine regulation that the biological drift method can highlight within the reference thresholds represented by the red lines in Figure 2. By visualizing these individual trajectories, we can observe gradual oscillations or trends that may reflect an underlying evolution of the patient’s overall biological state. This dynamic perspective contrasts with a pointwise interpretation based solely on thresholds, thereby enabling closer attention to fluctuations occurring within reference ranges.

Importantly, such intra-threshold variations often precede the crossing of reference limits, suggesting that certain biological parameters begin to drift before reaching overtly abnormal values. Monitoring these dynamics, particularly through the analysis of drift zones, provides a way to follow the patient’s trajectory over time. In our albumin example, this approach reveals a gradual transition, with an initial Opt phase at the beginning of 2014, followed by an LD phase in mid-2019 and a VLD phase in 2022, preceding hospitalization with surgery on June 15, 2022 (see red triangles in Figure 2).

### First hospitalization event survival analysis with Kaplan-Meier and log-rank tests

#### Univariate analysis

First, we used the Kaplan-Meier estimator to estimate survival curves up to the first hospitalization event. In this study, 58% of patients experienced at least one hospitalization, while the remaining 42% without any event were considered right-censored.

Thirty-two biological parameters were significant in the global log-rank test (see Supplementary Table 1). Among these 32 markers, 14 were significant for at least one class (including ARI and BRI). Nine were significant only for BRI or ARI: Neutrophils (%)-BRI (corrected p-value=0.040), Platelets-ARI (corrected p-value=0.003), C-reactive protein-ARI (corrected p-value=0.036), fecal calprotectin-ARI (corrected p-value=0.003), Lactate Dehydrogenase-ARI (corrected p-value=0.032), Free T4-BRI (corrected p-value<0.001), Calcium-BRI (corrected p-value=0.003), Iron-BRI (corrected p-value=0.036), and Total cholesterol-BRI (corrected p-value=0.006). Five markers showed significant within-reference drift classes (excluding ARI and BRI): Lymphocytes (%)-HD, Monocytes (%)-HD, aspartate aminotransferase (AST)-VHD, Albumin-HD and -VHD, and Potassium-HD; Albumin-BRI was also significant (see Supplementary Table 2).

For example, for albumin, Figure 3 shows the survival analysis results stratified by drift level. A global log-rank test yielded a *p*-value < 0.0001, indicating the presence of at least one significant difference between drift levels. This difference is reflected in distinct survival probabilities, with reduced survival observed when albumin values are low (BRI and VLD) compared to the biological optimum (Opt). Pairwise comparisons revealed statistically significant differences between VLD and Opt (*p*-value = 0.033), BRI and Opt (*p*-value < 0.0001), and BRI and VLD (*p*-value = 0.0032). The risk of a first hospitalization event increases as albumin levels decline, even before they fall below the lower reference threshold.

**Fig. 3.**
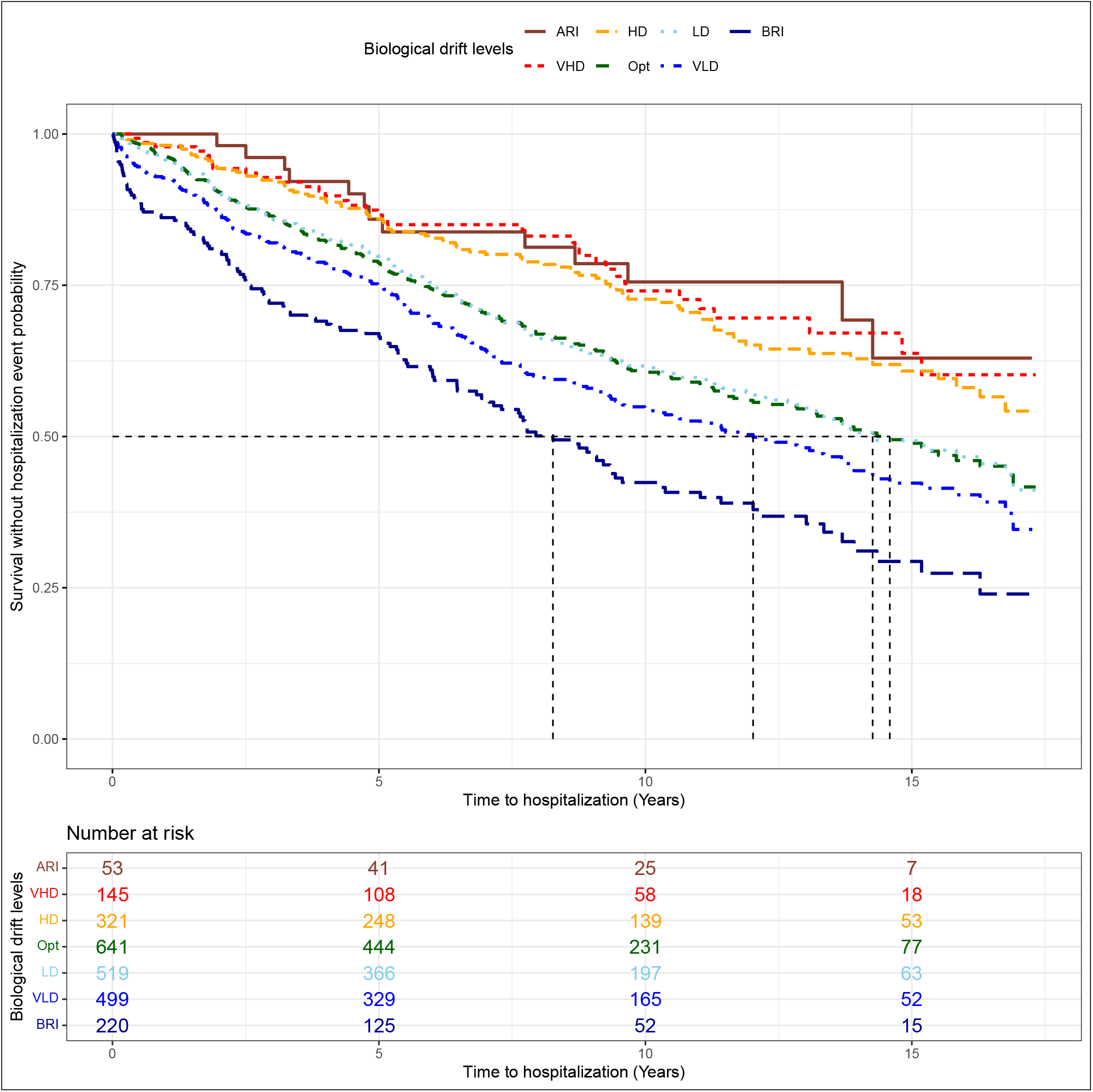
Albumin: estimated Kaplan-Meier survival curves by drift level at the time of the first hospitalization event.

#### Bivariate analysis with CRP

Still using Kaplan-Meier models with log-rank analyses to assess the risk of a first hospitalization event, we then simultaneously stratified each parameter by CRP status: CRP *≤* 5, considered normal (Opt), and CRP *>* 5, considered elevated (ARI), in line with clinical practice. We considered the CRP value measured on the same date as the other biological parameters tested. Combination classes were compared using pairwise log-rank tests (see Supplementary Table 3).

For example, for albumin, the inclusion of an additional variable such as CRP allowed finer stratification than before and highlighted steeper declines in survival probability when CRP *>* 5 was combined with low albumin levels (BRI or VLD) in Figure 4 (b). A global log-rank test again yielded a *p*-value < 0.0001. Albumin BRI remained significant regardless of CRP levels. Moreover, when CRP values exceeded 5 and were combined with albumin VLD, the association became statistically significant, underscoring the need to account for the combined effect of these parameters, even though albumin levels were still within the reference interval.

**Fig. 4.**
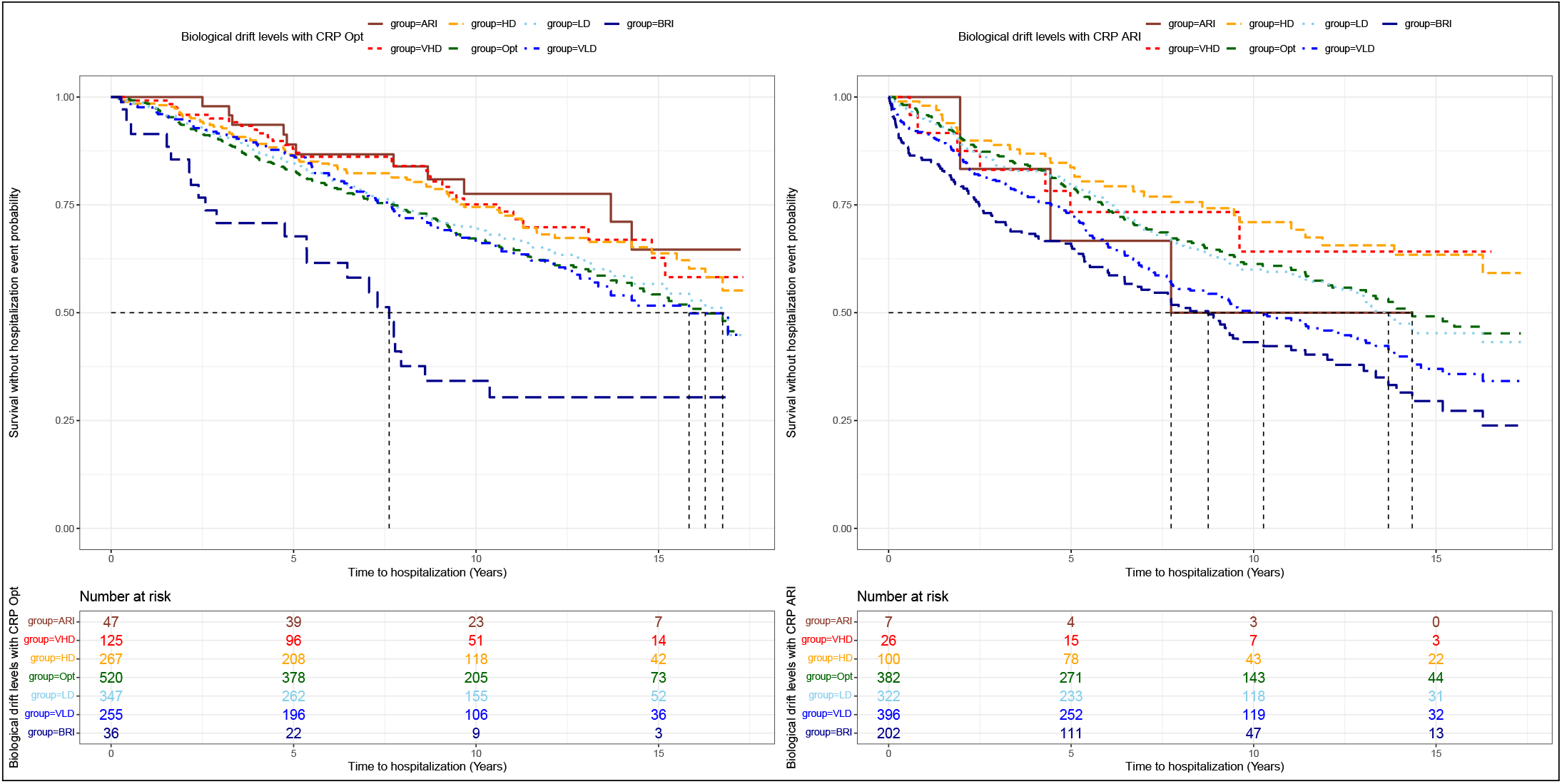
Albumin: estimated Kaplan-Meier survival curves by drift level combined with CRP ≤ 5 (a) or > 5 (b) at the time of the first hospitalization event.

### Recurrent event analysis with Cox models

Cox models were run for parameters whose drift levels (excluding ARI and BRI) were significantly different from Opt in Kaplan-Meier analyses: Lymphocytes (%), Monocytes (%), aspartate aminotransferase (AST), Albumin, and Potassium.

#### Analysis with biological drift levels

In the PWP-GT model with frailty, significantly higher risks were observed for AST-LD (HR=1.29, p-value=0.029), AST-BRI (HR=1.81, p-value<0.001), Albumin-VLD (HR=1.41, p-value=0.008), CRP-ARI (HR=1.69, p-value<0.001), and Monocytes (%)-VLD (HR=1.33, p-value=0.015); see Figure 5. Other combinations, such as Lymphocytes (%)-VLD, were significant in the PWP-TT model with frailty (HR=1.28, p-value=0.043) and in the AG model (HR=1.26, p-value=0.019), while Monocytes (%)-LD was significant in the PWP-GT model (HR=1.26, p-value=0.018); see Supplementary Figure 2.

**Fig. 5.**
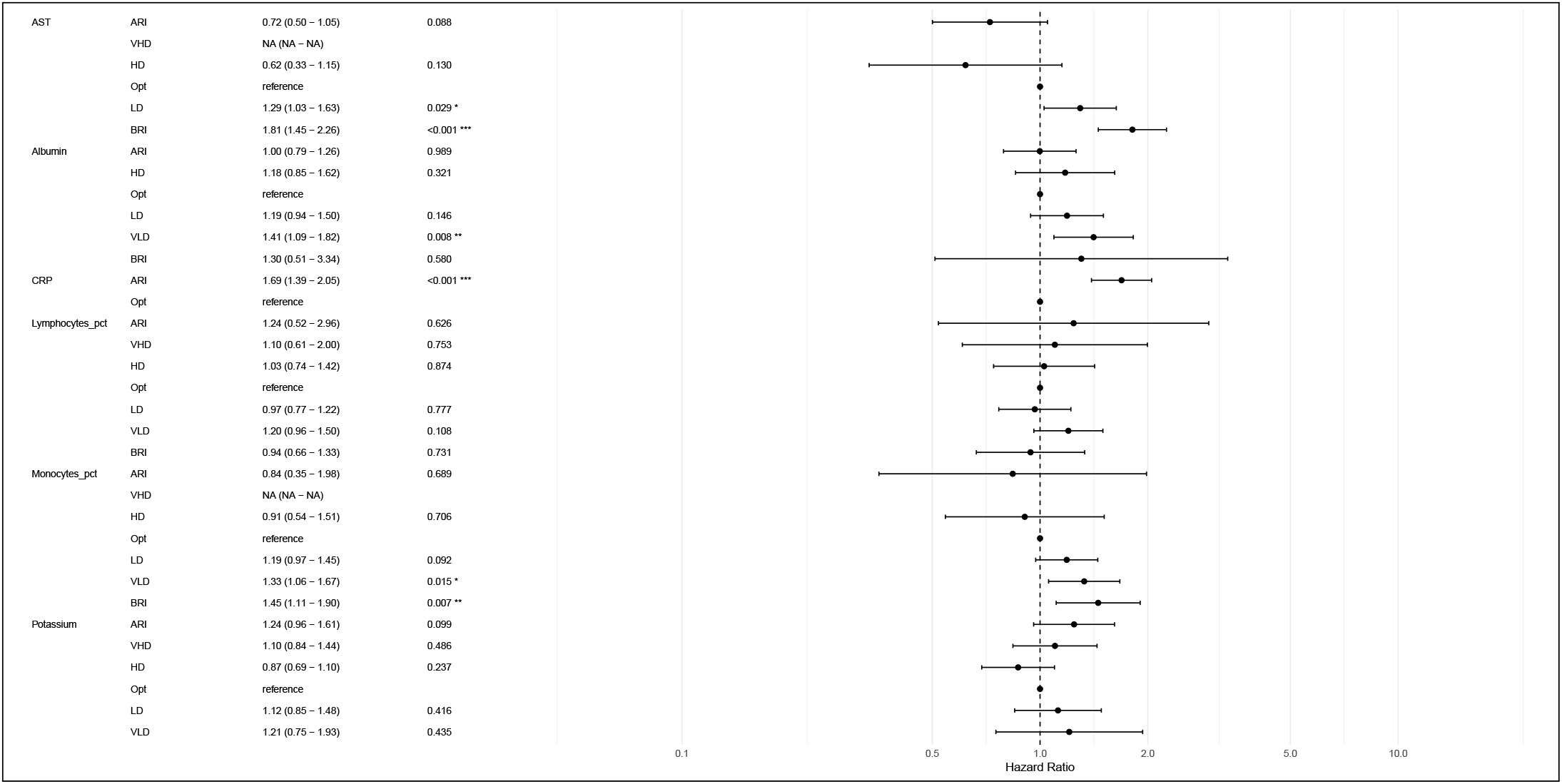
Hazard ratios estimated with the PWP-GT model with frailty.

#### Analysis with conventional reference intervals

To compare with standard practice, Cox models were run using a three-class stratification (BRI, WRI, and ARI), with WRI encompassing VHD, HD, Opt, LD, and VLD. This was the “without drift levels” setting. In the PWP-GT model with frailty, Monocytes (%) BRI had an HR of 1.28 (1.01-1.63) and a p-value of 0.041* without drift levels, and an HR of 1.45 (1.11-1.90) and a p-value of 0.007** with drift levels; see Supplementary Figures 3 and 4 for comparison with Figure 5. The Opt group in the drift-level setting is therefore at lower risk than the Opt group in the setting without drift levels, where all within-reference-interval values are pooled.

### Interaction between biological drift and hospitalization type

When risk was computed separately for hospitalizations without surgery and with surgery, some significant associations were observed (especially for the PWP-GT model with frailty): Albumin-LD without surgery (HR=1.40, p-value=0.037) and Potassium-ARI with surgery (HR=1.73, p-value=0.007). Overall, biological drift levels were more often associated with the risk of hospitalizations requiring surgery (see Figure 6).

**Fig. 6.**
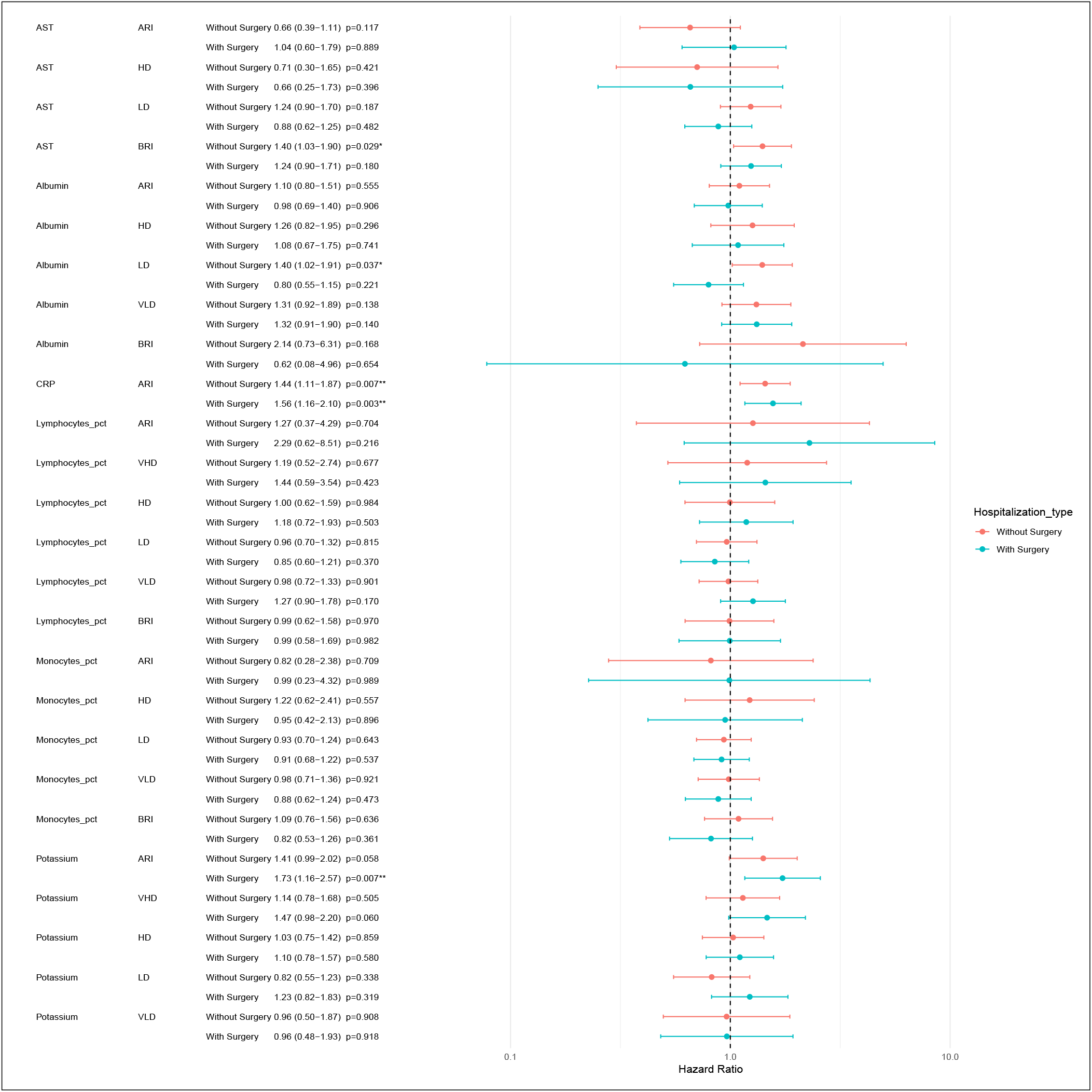
Hazard ratios estimated with the PWP-GT model with frailty, according to hospitalization type.

Among the significant associations in the PWP-GT model with frailty, the AST-BRI combination was more strongly associated with hospitalization without surgery (HR=1.40, p-value=0.029). Albumin-VLD showed similar associations with hospitalization without and with surgery (HR=1.31, p-value=0.138 and HR=1.32, p-value=0.140, respectively), as did CRP-ARI (HR=1.44, p-value=0.007 and HR=1.56, p-value=0.003, respectively); see Figure 6. See Supplementary Figure 5 for the results of all models.

## Discussion

In Kaplan-Meier models with log-rank analyses, low Neutrophils (%), high Platelets, high C-reactive protein, high fecal calprotectin, high Lactate dehydrogenase, low Free T4, low Calcium, low Iron, and low Total cholesterol were associated with a significant change in the risk of a first hospitalization event (higher or lower risk than Opt), but only for values in classes above or below the reference intervals. Biological drift levels for Lymphocytes (%), Monocytes (%), aspartate aminotransferase (AST), Albumin, and Potassium were found to be linked to hospitalization risk (first hospitalization event in KM or recurrent events in Cox models).

The comparison of Cox models using drift levels and conventional reference intervals highlighted the lower-risk profile of the Opt drift class compared with the simple withinreference-interval classification, especially for Monocytes (%). This suggests that these reference populations are biologically relevant for biomarker stratification.

The relative decrease in lymphocytes (%) and monocytes (%) observed in our cohort should be interpreted cautiously, as these variations remained within normal ranges and likely reflect a broader shift in leukocyte distribution rather than lineage-specific deficits. Increasing evidence suggests that composite inflammatory indices, particularly the neutrophil-to-lymphocyte ratio, are more robust markers of disease activity and prognosis in Crohn’s disease than absolute cell counts (22, 23). In this context, relative lymphopenia and monocytopenia may primarily indicate a proportional expansion of neutrophils, consistent with sustained activation of innate immunity. Neutrophils play a central role in intestinal inflammation through cytokine release, oxidative stress, and tissue infiltration (24). Additionally, lymphocyte redistribution toward inflamed mucosal sites may contribute to lower circulating levels (25), while monocyte trafficking to the gut and differentiation into pro-inflammatory macrophages is well established (26). Altogether, these findings support the interpretation of our results as reflecting a neutrophil-dominant systemic inflammatory state rather than isolated alterations in lymphocyte or monocyte compartments.

Lower albumin levels, even within the normal range, emerged as a relevant marker of adverse outcomes. Hypoalbuminemia in inflammatory bowel disease reflects a combination of systemic inflammation, reduced hepatic synthesis, increased intestinal loss, and heightened catabolism (27). It is also closely linked to impaired nutritional status and sarcopenia, both of which are highly prevalent in Crohn’s disease and associated with poorer clinical outcomes (28, 29). Importantly, albumin has direct pharmacokinetic implications: lower levels are associated with increased clearance of monoclonal antibodies such as infliximab, leading to reduced drug exposure and diminished therapeutic response (30, 31). Our findings therefore support a continuous, rather than threshold-based, interpretation of albumin, highlighting its integrative role as a marker of inflammatory burden, nutritional status, and drug disposition.

The association between lower AST levels and increased relapse risk may appear counterintuitive, as clinical attention typically focuses on elevated transaminases. However, AST is not liver-specific and is also expressed in skeletal muscle, where it reflects cellular turnover and metabolic activity. Lower AST levels have been associated with reduced muscle mass and sarcopenia in several clinical settings (32, 33). In Crohn’s disease, sarcopenia is increasingly recognized as a key component of disease burden, linked to chronic inflammation, malnutrition, and worse outcomes (28). Within this framework, low AST may act as an indirect biomarker of diminished muscle reserve and systemic frailty. This interpretation is consistent with the broader pattern observed in our study, in which markers reflecting a catabolic state and altered body composition were associated with poorer prognosis.

Higher potassium levels were specifically associated with an increased risk of hospitalization requiring surgery, but not with overall or non-surgical admissions. This pattern suggests that potassium may reflect advanced or complicated disease rather than inflammatory activity alone. Mild elevations in potassium can occur in the context of systemic catabolism and cellular breakdown, with intracellular potassium release, particularly in patients with inflammation-driven muscle wasting (27, 28). In addition, subtle alterations in renal handling related to dehydration, inflammation, or medication exposure may contribute to values in the upper normal range (34). Given the tight physiological regulation of potassium, such variations may indicate early homeostatic stress and systemic fragility. The absence of an association with non-surgical hospitalization supports its role as a marker of disease complexity rather than flare severity alone. However, these findings should be interpreted cautiously due to potential technical confounding factors, such as false hyperkalemia.

When stratifying hospitalizations according to surgical status (with vs. without surgery), most variables exhibited a consistent risk pattern across both types of events. However, notable differences emerged for albumin levels. Specifically, low albumin values were more strongly associated with hospitalizations without surgery. This trend was also observed when using the BRI classification, although it did not reach statistical significance in either analysis. This differential association may reflect the underlying pathophysiological role of hypoalbuminemia in Crohn’s disease. Low albumin levels are likely to represent a composite marker of systemic inflammation, increased intestinal protein loss, and impaired nutritional status. As such, hypoalbuminemia may be more closely linked to acute inflammatory disease activity requiring medical management and supportive care than to structural complications such as strictures or fistulas, which more frequently lead to surgical intervention. This suggests that albumin primarily captures the burden of inflammatory and catabolic processes, which may preferentially drive nonsurgical hospitalizations.

This study highlights the value of personalized reference ranges and the potential of biological drift to predict poor outcomes in Crohn’s disease within conventional reference intervals. In line with the growing interest in predictive biomarkers, our findings support a novel focus on subclinical changes, where risk accumulates progressively rather than appearing only when conventional thresholds are crossed.

Among the study limitations, we observed variability in both biomarker measurement frequency and patient profiles. Some parameters were less frequently assessed, reducing statistical power and limiting their contribution to the models. Similarly, Albumin-BRI did not remain significant in Cox models (whereas the VLD class did), which may reflect measurement variability or a smaller sample size in this specific stratum. Regarding study design, patients differed widely in disease phenotype, treatment exposure, and clinical trajectory. Treatments, in particular, can influence biomarker dynamics, yet they were not explicitly modeled here due to their diversity, the absence of precise dosing information, and the incompleteness of some treatment records. This variability likely contributes to residual confounding, and more targeted analyses focusing on specific drug classes could provide valuable additional insights.

For future work, it would be interesting to study Neutrophils (%), Platelets, C-reactive protein, fecal calprotectin, Lactate dehydrogenase, Free T4, Calcium, Iron, and Total cholesterol in Cox models to assess whether values above or below reference intervals are associated with a lower or higher risk of hospitalization. More sophisticated strategies, such as joint models combining longitudinal biomarker trajectories with recurrent events, could in principle provide a deeper understanding of the dynamic interplay between biomarkers, treatments, and outcomes. In addition, multidimensional or topological approaches such as Datascape (35) or GEDO (36), both developed by our team, may help characterize global patient profiles beyond the analysis of individual biological parameters.

Beyond Crohn’s disease, monitoring biological drift could prove valuable across a wide range of conditions, particularly chronic diseases that may manifest silently or with overt clinical activity and whose temporal evolution is often complex, marked by phases of treatment, exacerbation, remission, or diagnostic transition. By capturing these subtle and continuous biological variations in commonly measured biomarkers, this approach paves the way for a more dynamic and adaptive model of patient monitoring, better aligned with the heterogeneous and evolving nature of human health trajectories.

## Conclusion

This study highlights the value of routinely measured blood biomarkers for monitoring the risk of rehospitalization in patients with Crohn’s disease. Abnormalities defined according to conventional reference intervals showed prognostic value, reflecting different dimensions of hospitalization risk for elevated CRP, high Potassium, low AST, and low Monocytes (%), for instance. Beyond these established signals, the incorporation of biological drift revealed that even fluctuations within conventional reference intervals are clinically relevant for Albumin, Monocytes (%), and Lymphocytes (%), showing a progressive increase in hazard ratios across drift categories and indicating that risk accumulates gradually rather than only at pathological extremes. These alterations can be explained by neutrophil imbalance, nutritional status, sarcopenia, or treatment-related sources of variation.

By capturing these early signals, biological drift provides a complementary dimension to conventional reference intervals, improving risk stratification and offering a more dynamic picture of disease evolution. Integrating such markers and their variations within normal ranges into routine monitoring could therefore help anticipate hospitalizations and guide more personalized management strategies in Crohn’s disease.

## Data Availability

All data produced in the present work are contained in the manuscript.

## Contributors

AH conducted the methodological work, performed the analyses, and drafted the manuscript. JR contributed to the scientific design and manuscript writing. CB contributed to manuscript writing. LE contributed to the scientific design and coordinated the project. NM initiated the project, extracted the data, and contributed to the scientific design, particularly regarding statistical methods. MT performed data extraction and cleaning. RB contributed to the scientific design and project coordination. EL provided scientific leadership for the project, interpreted the results, and contributed to manuscript writing.

## Acknowledgments

We thank all patients included in this study, the staff at Liège University Hospital for data management, and all gastroenterologists for patient follow-up and inclusion in the database.

## Funding

NA.

## Competing interests

RB is the founder of Bio Logbook, and this did not affect the design, data collection, analysis, conclusions, decision to publish, or preparation of the manuscript. All other authors declare no potential conflicts of interest.

## Notes

### Author Declarations

Ethics committee of University Hospital of Liège gave ethical approval for this work

