## Supplementary Material for "Biological drifts within normal ranges allow the detection of Crohn’s disease patients at high risk of rehospitalization"

### Supplementary Information

#### A. Optimized and personalized reference populations.

The dataset from the Montpellier community laboratory included all individuals aged $\geq 18$ years who underwent testing between October 2016 and December 2019, comprising 996,975 distinct individuals (42.2% male, 57.8% female) with a mean age of 54.5 ± 19.9 years (range: 18–110 years) and a total of 37,677,310 test results. Each patient's laboratory report included, at a minimum, red blood cell (RBC) count, hemoglobin, hematocrit, mean corpuscular volume (MCV), mean corpuscular hemoglobin (MCH), mean corpuscular hemoglobin concentration (MCHC), platelet count, and white blood cell (WBC) count, including total leukocytes, lymphocytes, monocytes, neutrophils, eosinophils, and basophils. Preanalytical and analytical details are provided in (1).

#### B. Recurrence of hospitalization events.

Recurrence was defined as the occurrence of two or more hospitalizations during the observation period. A more detailed representation of event occurrence over time is shown in Supplementary Figure 1 for 30 patients randomly selected from the cohort. Substantial heterogeneity in follow-up duration was observed, ranging from a few years to more than 15 years (between 2005 and 2023). In addition, the frequency, number, and nature of hospitalizations varied considerably, illustrating the variability and complexity of the disease. Supplementary Figure 1(b) and (c) show the mean cumulative function (MCF), which estimates the mean number of events per patient over time, based on the entire cohort described in Table 1. Supplementary Figure 1(c) compares the MCF by sex; no major difference was observed between males and females, indicating that the average rate of hospitalization was comparable between the two groups. Finally, although no global pattern emerged, individual visualizations highlighted substantial interindividual heterogeneity, underlining the diversity of clinical trajectories within the disease. This variability also extends to the collection of biological data: blood samples and other biomarker measurements were performed at irregular and often unbalanced intervals across patients, creating specific challenges for their management.

#### C. Survival Modeling.

***C.1. Model Framework.*** The analysis of real-world data from a hospital setting presents specific methodological challenges, particularly because of the irregular and unbalanced nature of biological measurements. Biomarkers may be assessed at variable intervals, often driven by the patient's clinical condition, which introduces potential bias in their interpretation. In this context, survival methods provide an appropriate framework for modeling the time to a clinical event of interest, such as hospitalization, while accounting for the structure of longitudinal data.

A common challenge lies in the handling of censoring, as not all patients necessarily experience the event of interest during the follow-up period. These patients are said to be right-censored: their follow-up time is known, but the event has not occurred by the end of the observation period and may still occur after the study cutoff date (2023 in the case of this study). For each individual $i$, let $T_i$ denote the event time and $C_i$ the censoring time. We assume independent censoring, i.e. $C_i \perp T_i$, in the absence of evidence to suggest otherwise. In recurrent-event settings, a subject $i$ is considered at risk at time $t$ if he or she remains under observation in the study ($C_i \geq t$). Depending on the modeling framework, the subject may remain at risk after experiencing previous events. Let $Y_i(t)$, for all $i \in \{1, \ldots, N\}$, denote the at-risk indicator function, equal to 1 if subject $i$ is under observation at time $t$. It is crucial to properly account for censored and recurrent observations in the modeling to avoid estimation bias.

In this context, the use of survival models makes it possible to analyze the occurrence of events over time while accounting for censoring, and to explore how biomarker values may be associated with Crohn's disease rehospitalization.

Several models were considered: first, a simple non-parametric model based on the first event (Kaplan-Meier); second, a parametric model accounting for independent recurrence between events (Andersen–Gill); and finally, a model accounting for dependence on event order (Prentice, Williams & Peterson).

**C.2. Kaplan–Meier estimator.** The Kaplan–Meier estimator provides a non-parametric estimate of the survival function based on the ordered event times (2). To assess differences between groups, survival curves can then be statistically compared using the log-rank test (3). One approach is to consider only the first event occurring for each patient. This strategy simplifies the model by reducing each individual to a single time-to-event observation. Thus, patients with multiple successive hospitalizations are represented only by the first.

**C.3. Cox model.** In this study, the recurrent nature of hospitalizations must be considered; Cox regression models (4) are well suited to modeling multiple events within the same individual. The Cox model estimates the instantaneous hazard of an event as a function of explanatory covariates according to:

$$h(t \mid \mathbf{X}) = h_0(t) \cdot \exp(\boldsymbol{\beta}^\top \mathbf{X})$$

where $h(t \mid \mathbf{X})$ is the hazard function at time $t$, $h_0(t)$ is the baseline hazard function, $\mathbf{X}$ is the covariate vector, and $\boldsymbol{\beta}$ is the vector of coefficients.

**C.4. Andersen-Gill model.** The Andersen–Gill (AG) model (5) extends the Cox model to recurrent events by treating all hospitalizations as a single counting process. This first approach to recurrent hospitalizations can be seen as a simplified model, as it does not explicitly account for each patient's individual history. The sequence of events is not modeled as a dependent process, which is equivalent to assuming that the number of prior hospitalizations does not directly influence the conditional hazard function.

**C.5. Prentice, Williams & Peterson model.** To account for the dependence between successive hospitalizations as a function of their order, the Prentice, Williams & Peterson (PWP) model (6) conditions the occurrence of the $k^{\text{th}}$ event on that of the $(k-1)^{\text{th}}$ event. This model is based on stratification by event order, allowing each recurrent event to have its own baseline hazard function. Two variants of the model exist, depending on how time is defined: *Total Time* (time since study entry) and *Gap Time* (time since the previous event). The choice of the appropriate variant is discussed later.

**C.6. Frailty integration.** To model unobserved heterogeneity among patients, a multiplicative random effect known as frailty (7) can be introduced into the Cox model. This approach captures individual-level differences not explained by measured covariates, assuming that each individual has a specific risk factor. The Cox model with frailty is defined as:

$$h(t \mid \mathbf{X}, u) = u \cdot h_0(t) \cdot \exp\left(\boldsymbol{\beta}^\top \mathbf{X}\right)$$

where $u$ is the frailty component, generally modeled as a positive random variable following a gamma distribution, reflecting the degree of inter-individual variability.

**C.7. Association between surgery and drift levels.** Finally, to assess whether the association between drift levels and hospitalization differs according to the presence or absence of surgery, we introduced an interaction term between drift levels and surgery status in the survival models presented above. This approach allows us to estimate risk factors for each drift level separately for surgical and non-surgical hospitalizations.

### Supplementary Figures

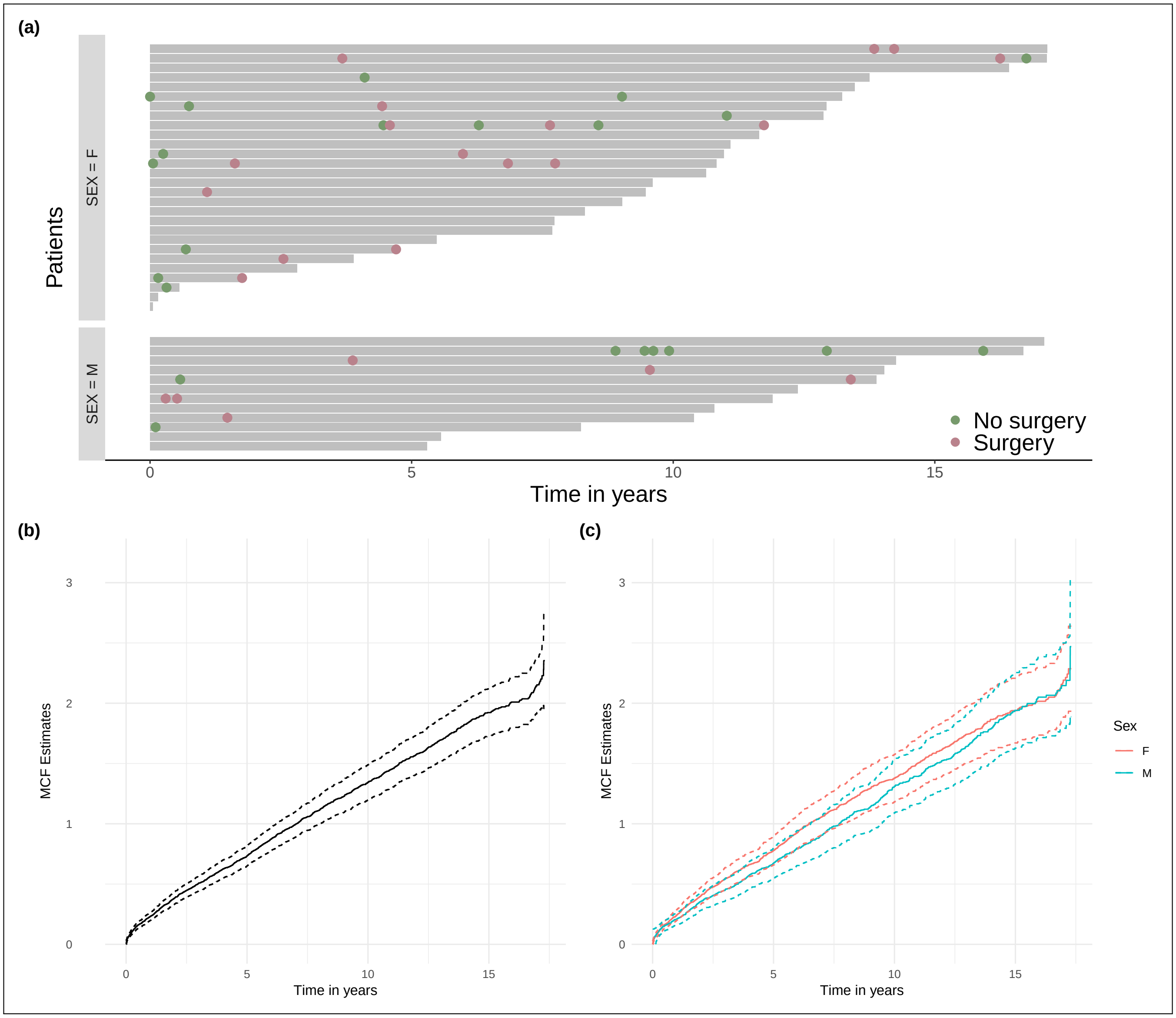


**Fig. 1.** Recurrence of hospitalization

(a) Hospitalizations over time, with or without surgery. Each line represents one individual, stratified by sex. Thirty patients were randomly selected. (b) Mean cumulative function (MCF), which estimates the average number of events per patient over time with a 95% confidence interval. (c) Same as (b), stratified by sex.

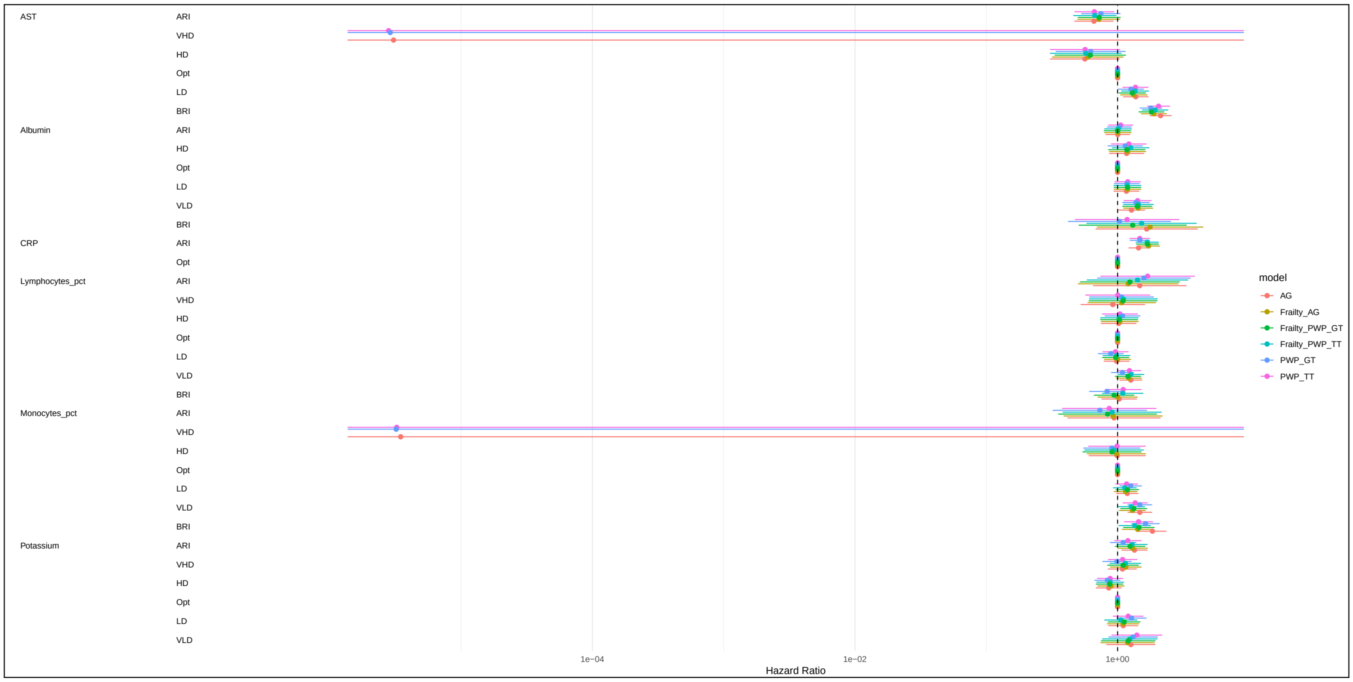


**Fig. 2.** Hazard ratios estimated using all Cox models, with drift levels.

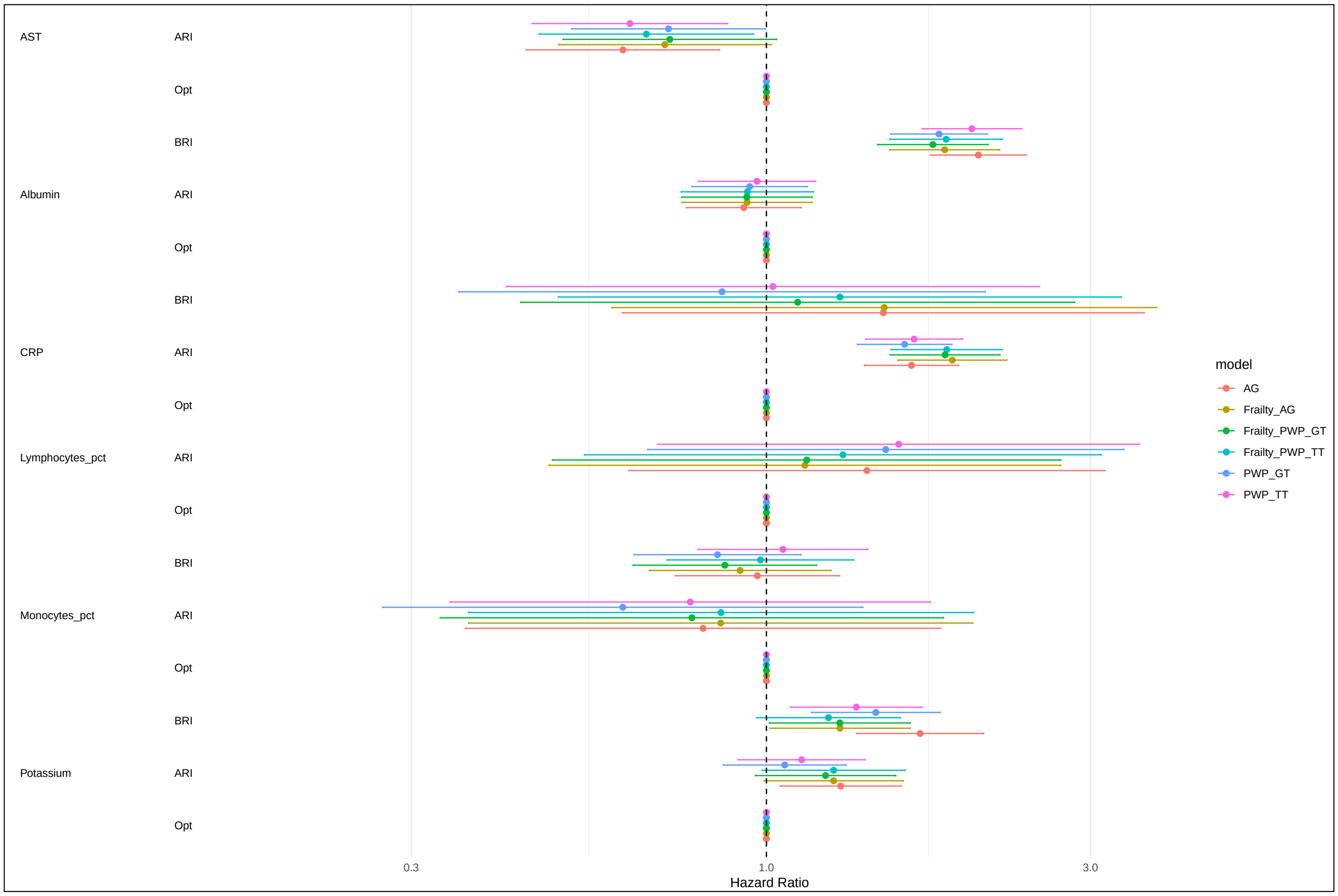


**Fig. 3.** Hazard ratios estimated using all Cox models, without drift levels.

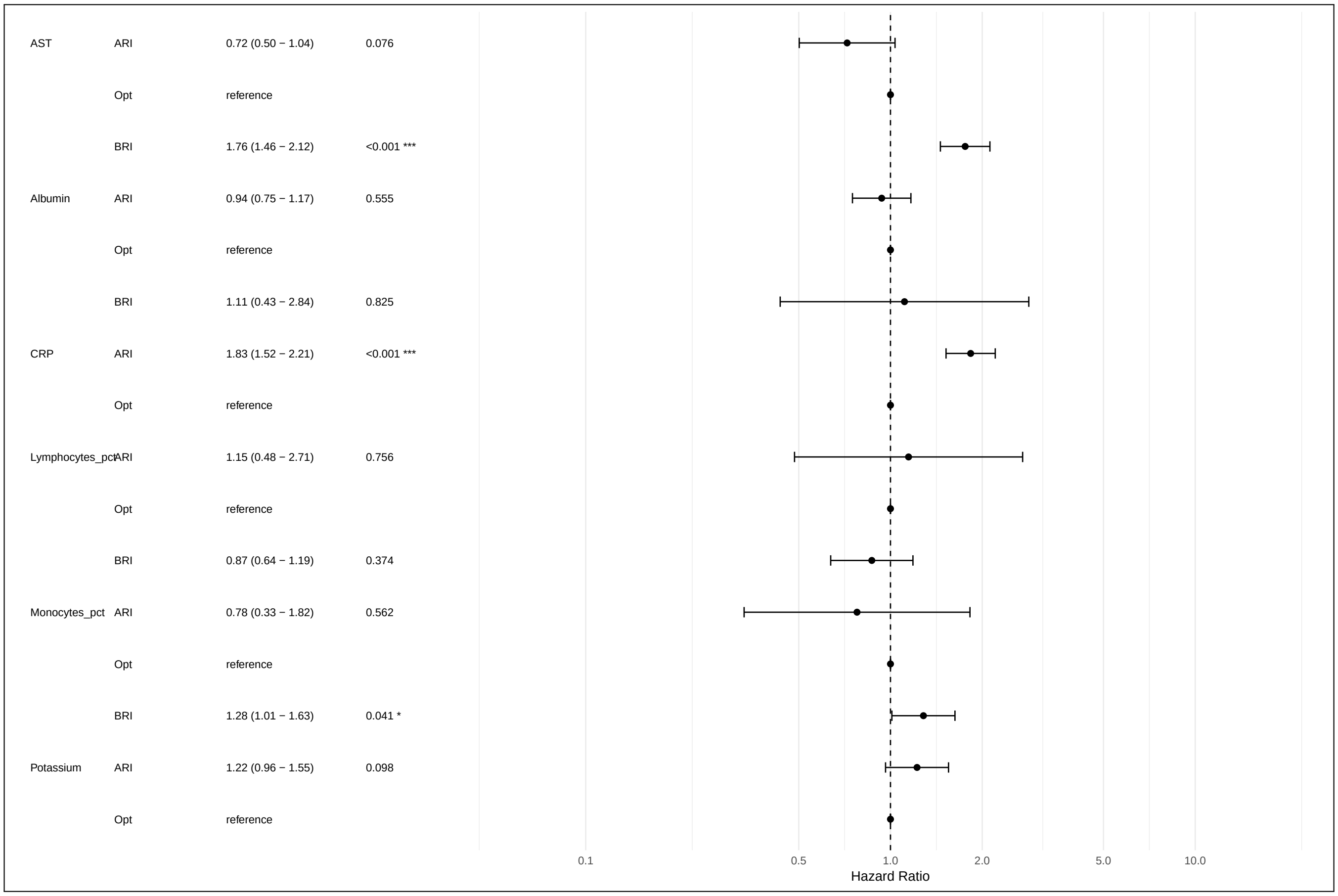


**Fig. 4.** Hazard ratios estimated using the PWP-GT model with frailty, without drift levels.

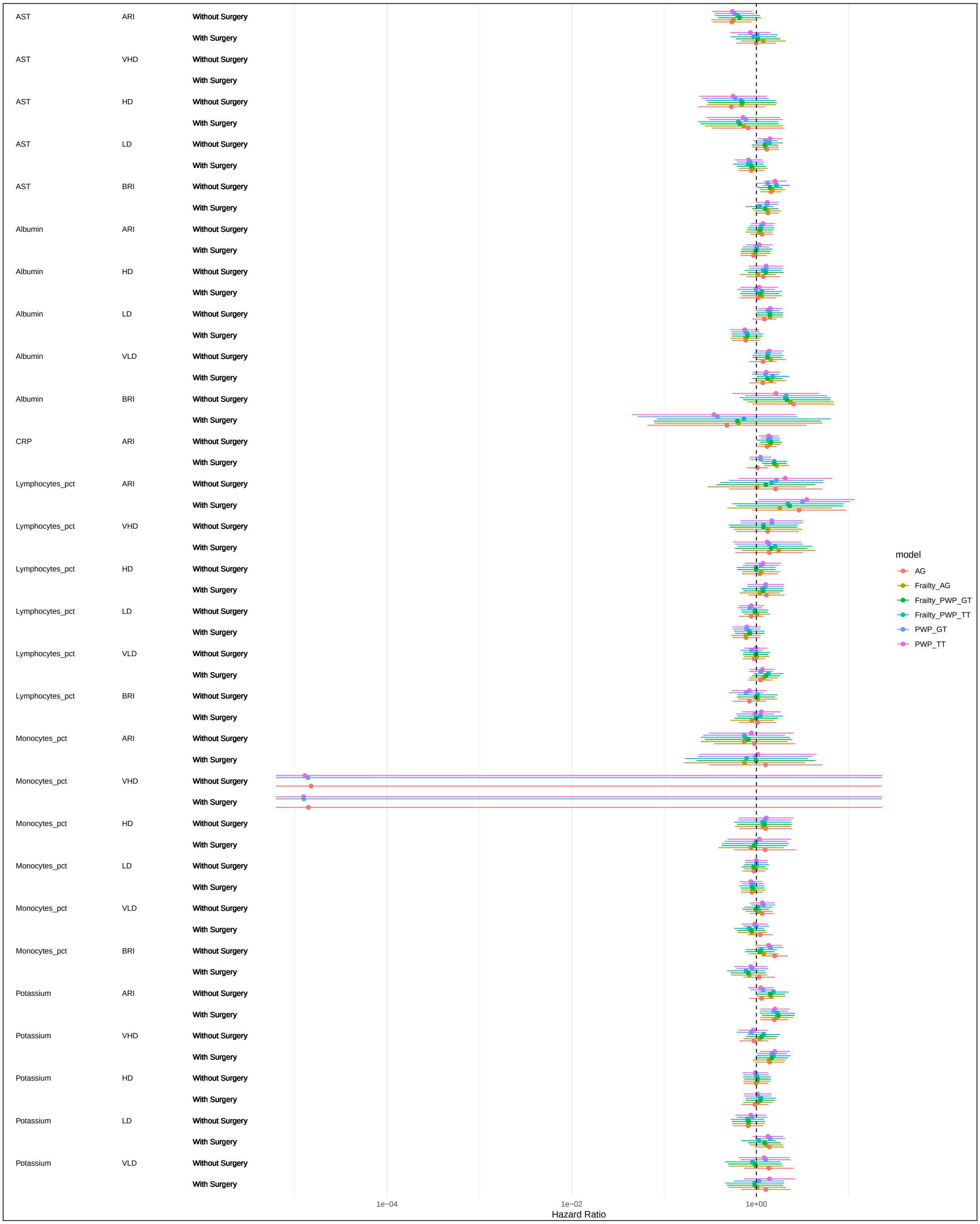


**Fig. 5.** Hazard ratios for drift classes estimated using all Cox models, according to hospitalization type.

### Supplementary Tables

**Table 1:** Global log-rank tests of the 57 parameters tested (ordered by p-value). BH-corrected p-value: p-value corrected using the Benjamini-Hochberg method.

| Parameter | p-value | BH-corrected p-value |
|---|---|---|
| Albumin | 1.67e-15 | 9.49e-14 |
| Lymphocytes (pct) | 6.06e-11 | 1.73e-09 |
| Calcium | 3.37e-08 | 5.43e-07 |
| Iron | 3.81e-08 | 5.43e-07 |
| Neutrophils (pct) | 2.14e-07 | 2.44e-06 |
| Total cholesterol | 5.31e-07 | 5.05e-06 |
| Free T4 | 1.50e-06 | 1.22e-05 |
| AST | 1.78e-06 | 1.26e-05 |
| Potassium | 2.97e-06 | 1.88e-05 |
| Neutrophils | 6.47e-05 | 3.69e-04 |
| Hemoglobin | 8.19e-05 | 4.24e-04 |
| Fecal calprotectin | 1.12e-04 | 5.31e-04 |
| Platelets | 1.29e-04 | 5.65e-04 |
| Total proteins | 2.46e-04 | 9.34e-04 |
| LDH | 2.32e-04 | 9.34e-04 |
| ESR | 8.46e-04 | 3.01e-03 |
| Hematocrit | 1.34e-03 | 4.49e-03 |
| Fibrinogen | 1.42e-03 | 4.49e-03 |
| Monocytes (pct) | 2.69e-03 | 8.06e-03 |
| C-reactive protein | 2.98e-03 | 8.08e-03 |
| Chloride | 2.89e-03 | 8.08e-03 |
| LDL cholesterol | 3.49e-03 | 9.05e-03 |
| Eosinophils (pct) | 3.77e-03 | 9.06e-03 |
| Transferrin | 3.81e-03 | 9.06e-03 |
| Basophils (pct) | 4.42e-03 | 0.0101 |
| Total lymphocytes | 7.88e-03 | 0.0173 |
| Urea | 0.011 | 0.0233 |
| Sodium | 0.0148 | 0.03 |
| Lipase | 0.0186 | 0.0366 |
| ALT | 0.0204 | 0.0375 |
| Triglycerides | 0.0202 | 0.0375 |
| Bilirubin | 0.0234 | 0.0418 |
| Red blood cells | 0.0428 | 0.074 |
| Creatine kinase | 0.0461 | 0.0772 |
| White blood cells | 0.0534 | 0.0869 |
| Bicarbonate | 0.0585 | 0.0926 |
| Vitamin D | 0.0659 | 0.102 |
| Eosinophils | 0.0684 | 0.103 |
| MCHC | 0.114 | 0.163 |
| Phosphate | 0.114 | 0.163 |
| Uric acid | 0.12 | 0.167 |
| Magnesium | 0.188 | 0.255 |
| Amylase | 0.222 | 0.294 |
| GGT | 0.235 | 0.305 |
| Ferritin | 0.289 | 0.366 |
| MCH | 0.34 | 0.422 |
| Basophils | 0.349 | 0.423 |
| Creatinine | 0.375 | 0.446 |
| Monocytes | 0.447 | 0.52 |
| MCV | 0.501 | 0.571 |
| Folate | 0.526 | 0.588 |
| Vitamin B12 | 0.616 | 0.675 |
| Alkaline phosphatase | 0.665 | 0.715 |
| Glucose | 0.86 | 0.908 |
| Reticulocytes | 0.883 | 0.915 |
| Reticulocytes (pct) | 0.932 | 0.949 |
| TSH | 0.979 | 0.979 |

**Table 2:** Pairwise log-rank tests of the parameters tested with the Opt class as the reference (ordered by parameter and drift class). BH-corrected p-value: p-value corrected using the Benjamini-Hochberg method.

| Parameter | Drift class | p-value | BH-corrected p-value |
|---|---|---|---|
| ALT | ARI | 0.135 | 0.307 |
| ALT | VHD | 0.182 | 0.368 |
| ALT | HD | 0.121 | 0.287 |
| ALT | LD | 0.804 | 0.939 |
| ALT | VLD | 0.823 | 0.939 |
| ALT | BRI | 0.929 | 0.954 |
| AST | ARI | 0.02 | 0.0974 |
| AST | VHD | 3.33e-04 | 6.92e-03 |
| AST | HD | 8.55e-03 | 0.0676 |

| Parameter | Drift class | p-value | BH-corrected p-value |
|---|---|---|---|
| AST | LD | 0.439 | 0.669 |
| AST | VLD | 0.439 | 0.669 |
| Albumin | ARI | 0.0345 | 0.143 |
| Albumin | VHD | 3.43e-03 | 0.0379 |
| Albumin | HD | 1.86e-03 | 0.028 |
| Albumin | LD | 0.919 | 0.954 |
| Albumin | VLD | 0.0331 | 0.143 |
| Albumin | BRI | 1.25e-06 | 1.16e-04 |
| Basophils (pct) | ARI | 0.086 | 0.237 |
| Basophils (pct) | VHD | 0.086 | 0.237 |
| Basophils (pct) | HD | 0.086 | 0.237 |
| Basophils (pct) | LD | 0.502 | 0.713 |
| Basophils (pct) | VLD | 0.418 | 0.648 |
| Basophils (pct) | BRI | 0.124 | 0.289 |
| Bilirubin | ARI | 0.217 | 0.419 |
| Bilirubin | VHD | 0.181 | 0.368 |
| Bilirubin | HD | 0.212 | 0.419 |
| Bilirubin | LD | 0.654 | 0.828 |
| Bilirubin | VLD | 0.654 | 0.828 |
| Bilirubin | BRI | 0.217 | 0.419 |
| C-reactive protein | ARI | 2.98e-03 | 0.0357 |
| Calcium | ARI | 0.0872 | 0.237 |
| Calcium | HD | 0.0205 | 0.0974 |
| Calcium | LD | 0.311 | 0.543 |
| Calcium | VLD | 0.0194 | 0.0974 |
| Calcium | BRI | 7.13e-05 | 2.96e-03 |
| Chloride | ARI | 0.0191 | 0.0974 |
| Chloride | VHD | 0.769 | 0.925 |
| Chloride | HD | 0.0491 | 0.173 |
| Chloride | LD | 0.182 | 0.368 |
| Chloride | VLD | 0.1 | 0.248 |
| Chloride | BRI | 0.972 | 0.983 |
| ESR | ARI | 0.0107 | 0.0774 |
| ESR | VHD | 0.0185 | 0.0974 |
| ESR | HD | 0.304 | 0.537 |
| ESR | BRI | 0.91 | 0.954 |
| Eosinophils (pct) | ARI | 0.0898 | 0.237 |
| Eosinophils (pct) | VHD | 0.0356 | 0.143 |
| Eosinophils (pct) | HD | 0.37 | 0.608 |
| Eosinophils (pct) | LD | 0.0607 | 0.206 |
| Eosinophils (pct) | VLD | 0.135 | 0.307 |
| Eosinophils (pct) | BRI | 0.341 | 0.584 |
| Fecal calprotectin | ARI | 1.12e-04 | 3.09e-03 |
| Fibrinogen | ARI | 0.0363 | 0.143 |
| Fibrinogen | VHD | 0.891 | 0.954 |
| Fibrinogen | HD | 0.855 | 0.953 |
| Fibrinogen | LD | 0.153 | 0.338 |
| Fibrinogen | VLD | 0.855 | 0.953 |
| Fibrinogen | BRI | 0.891 | 0.954 |
| Free T4 | ARI | 0.587 | 0.785 |
| Free T4 | VHD | 0.804 | 0.939 |
| Free T4 | HD | 0.986 | 0.987 |
| Free T4 | LD | 0.0114 | 0.0787 |
| Free T4 | BRI | 5.98e-06 | 3.31e-04 |
| Hematocrit | ARI | 0.511 | 0.72 |
| Hematocrit | VHD | 0.0972 | 0.248 |
| Hematocrit | HD | 0.167 | 0.355 |
| Hematocrit | LD | 0.897 | 0.954 |
| Hematocrit | VLD | 0.356 | 0.594 |
| Hematocrit | BRI | 0.0484 | 0.173 |
| Hemoglobin | ARI | 0.823 | 0.939 |
| Hemoglobin | VHD | 0.4 | 0.639 |
| Hemoglobin | HD | 0.0194 | 0.0974 |
| Hemoglobin | LD | 0.885 | 0.954 |
| Hemoglobin | VLD | 0.413 | 0.648 |
| Hemoglobin | BRI | 0.0356 | 0.143 |
| Iron | ARI | 0.105 | 0.257 |
| Iron | VHD | 0.0752 | 0.225 |
| Iron | HD | 0.0356 | 0.143 |
| Iron | LD | 0.793 | 0.939 |
| Iron | BRI | 3.01e-03 | 0.0357 |
| LDH | ARI | 2.33e-03 | 0.0322 |
| LDH | VHD | 0.596 | 0.785 |
| LDH | HD | 0.488 | 0.706 |
| LDH | LD | 0.596 | 0.785 |
| LDH | BRI | 0.285 | 0.514 |
| LDL cholesterol | ARI | 0.0893 | 0.237 |
| LDL cholesterol | VHD | 0.176 | 0.368 |
| LDL cholesterol | HD | 0.639 | 0.822 |
| LDL cholesterol | LD | 0.263 | 0.485 |
| LDL cholesterol | VLD | 0.0893 | 0.237 |

| Parameter | Drift class | p-value | BH-corrected p-value |
|---|---|---|---|
| Lipase | ARI | 0.889 | 0.954 |
| Lipase | HD | 0.489 | 0.706 |
| Lipase | LD | 0.826 | 0.939 |
| Lipase | VLD | 0.418 | 0.648 |
| Lipase | BRI | 0.0156 | 0.089 |
| Lymphocytes (pct) | ARI | 7.37e-03 | 0.0644 |
| Lymphocytes (pct) | HD | 1.40e-06 | 1.16e-04 |
| Lymphocytes (pct) | LD | 0.667 | 0.838 |
| Lymphocytes (pct) | BRI | 0.0252 | 0.116 |
| Monocytes (pct) | ARI | 0.189 | 0.377 |
| Monocytes (pct) | HD | 1.02e-03 | 0.017 |
| Monocytes (pct) | LD | 0.931 | 0.954 |
| Monocytes (pct) | VLD | 0.56 | 0.775 |
| Monocytes (pct) | BRI | 0.931 | 0.954 |
| Neutrophils | ARI | 0.165 | 0.355 |
| Neutrophils | VHD | 0.316 | 0.547 |
| Neutrophils | HD | 0.47 | 0.706 |
| Neutrophils | LD | 0.0486 | 0.173 |
| Neutrophils | VLD | 0.0107 | 0.0774 |
| Neutrophils | BRI | 0.165 | 0.355 |
| Neutrophils (pct) | ARI | 0.115 | 0.276 |
| Neutrophils (pct) | VHD | 0.885 | 0.954 |
| Neutrophils (pct) | HD | 0.744 | 0.902 |
| Neutrophils (pct) | LD | 5.54e-03 | 0.0511 |
| Neutrophils (pct) | VLD | 5.54e-03 | 0.0511 |
| Neutrophils (pct) | BRI | 3.89e-03 | 0.0404 |
| Platelets | ARI | 1.04e-04 | 3.09e-03 |
| Platelets | VHD | 0.0124 | 0.0793 |
| Platelets | HD | 0.138 | 0.309 |
| Platelets | LD | 0.833 | 0.941 |
| Platelets | BRI | 0.714 | 0.885 |
| Potassium | ARI | 0.099 | 0.248 |
| Potassium | VHD | 0.0993 | 0.248 |
| Potassium | HD | 7.53e-04 | 0.0139 |
| Potassium | LD | 0.398 | 0.639 |
| Potassium | VLD | 0.57 | 0.777 |
| Potassium | BRI | 0.0436 | 0.164 |
| Sodium | ARI | 0.0153 | 0.089 |
| Sodium | VHD | 0.987 | 0.987 |
| Sodium | HD | 0.358 | 0.594 |
| Sodium | LD | 0.289 | 0.516 |
| Sodium | VLD | 0.358 | 0.594 |
| Sodium | BRI | 0.926 | 0.954 |
| Total cholesterol | ARI | 0.386 | 0.628 |
| Total cholesterol | HD | 0.227 | 0.429 |
| Total cholesterol | LD | 0.0397 | 0.153 |
| Total cholesterol | VLD | 0.0544 | 0.188 |
| Total cholesterol | BRI | 2.37e-04 | 5.63e-03 |
| Total lymphocytes | ARI | 0.0758 | 0.225 |
| Total lymphocytes | VHD | 0.0648 | 0.215 |
| Total lymphocytes | HD | 0.0758 | 0.225 |
| Total lymphocytes | LD | 0.815 | 0.939 |
| Total lymphocytes | VLD | 0.576 | 0.777 |
| Total lymphocytes | BRI | 0.619 | 0.803 |
| Total proteins | ARI | 0.915 | 0.954 |
| Total proteins | VHD | 0.222 | 0.424 |
| Total proteins | HD | 0.277 | 0.505 |
| Total proteins | LD | 0.538 | 0.75 |
| Total proteins | VLD | 0.805 | 0.939 |
| Total proteins | BRI | 0.0119 | 0.0792 |
| Transferrin | ARI | 0.493 | 0.706 |
| Transferrin | VHD | 0.606 | 0.792 |
| Transferrin | HD | 0.956 | 0.973 |
| Transferrin | LD | 0.256 | 0.478 |
| Transferrin | BRI | 8.04e-03 | 0.0668 |
| Triglycerides | ARI | 0.494 | 0.706 |
| Triglycerides | VHD | 0.576 | 0.777 |
| Triglycerides | HD | 0.739 | 0.902 |
| Triglycerides | LD | 0.732 | 0.9 |
| Triglycerides | VLD | 0.494 | 0.706 |
| Triglycerides | BRI | 0.0133 | 0.0819 |
| Urea | ARI | 0.0689 | 0.216 |
| Urea | VHD | 0.0689 | 0.216 |
| Urea | HD | 0.0689 | 0.216 |
| Urea | LD | 0.478 | 0.706 |
| Urea | BRI | 0.701 | 0.875 |

**Table 3:** Pairwise log-rank tests stratified by CRP class (ordered by parameter, CRP class, and drift class). BH-corrected p-value: p-value corrected using the Benjamini-Hochberg method.

| Parameter | CRP class | Drift class | p-value | BH-corrected p-value |
|---|---|---|---|---|
| ALT | ARI | ARI | 0.524 | 1 |
| ALT | Opt | ARI | 0.0858 | 0.783 |
| ALT | ARI | VHD | 0.866 | 1 |
| ALT | Opt | VHD | 0.171 | 0.931 |
| ALT | ARI | HD | 0.388 | 1 |
| ALT | Opt | HD | 0.0418 | 0.591 |
| ALT | ARI | LD | 0.459 | 1 |
| ALT | Opt | LD | 0.804 | 1 |
| ALT | ARI | VLD | 0.671 | 1 |
| ALT | Opt | VLD | 0.589 | 1 |
| ALT | ARI | BRI | 0.851 | 1 |
| ALT | Opt | BRI | 0.349 | 1 |
| AST | ARI | ARI | 0.411 | 1 |
| AST | Opt | ARI | 0.194 | 0.94 |
| AST | ARI | VHD | 0.107 | 0.808 |
| AST | Opt | VHD | 0.0269 | 0.515 |
| AST | ARI | HD | 0.31 | 1 |
| AST | Opt | HD | 0.0182 | 0.432 |
| AST | ARI | LD | 0.645 | 1 |
| AST | Opt | LD | 0.807 | 1 |
| AST | ARI | VLD | 0.84 | 1 |
| AST | Opt | VLD | 0.84 | 1 |
| Albumin | ARI | ARI | 0.81 | 1 |
| Albumin | Opt | ARI | 0.157 | 0.931 |
| Albumin | ARI | VHD | 0.637 | 1 |
| Albumin | Opt | VHD | 0.12 | 0.846 |
| Albumin | ARI | HD | 0.114 | 0.823 |
| Albumin | Opt | HD | 0.0823 | 0.783 |
| Albumin | ARI | LD | 0.796 | 1 |
| Albumin | Opt | LD | 0.735 | 1 |
| Albumin | ARI | VLD | 5.89e-03 | 0.247 |
| Albumin | Opt | VLD | 0.994 | 1 |
| Albumin | ARI | BRI | 2.17e-05 | 4.29e-03 |
| Albumin | Opt | BRI | 3.74e-04 | 0.0499 |
| Alkaline phosphatase | ARI | ARI | 0.395 | 1 |
| Alkaline phosphatase | Opt | ARI | 0.165 | 0.931 |
| Alkaline phosphatase | ARI | VHD | 0.598 | 1 |
| Alkaline phosphatase | Opt | VHD | 0.395 | 1 |
| Alkaline phosphatase | ARI | HD | 0.997 | 1 |
| Alkaline phosphatase | Opt | HD | 0.592 | 1 |
| Alkaline phosphatase | ARI | LD | 0.433 | 1 |
| Alkaline phosphatase | Opt | LD | 0.433 | 1 |
| Alkaline phosphatase | ARI | BRI | 0.718 | 1 |
| Alkaline phosphatase | Opt | BRI | 0.971 | 1 |
| Amylase | ARI | ARI | 0.487 | 1 |
| Amylase | Opt | ARI | 0.954 | 1 |
| Amylase | ARI | HD | 0.786 | 1 |
| Amylase | Opt | HD | 0.644 | 1 |
| Amylase | ARI | LD | 0.845 | 1 |
| Amylase | Opt | LD | 0.797 | 1 |
| Amylase | ARI | BRI | 0.773 | 1 |
| Amylase | Opt | BRI | 0.986 | 1 |
| Basophils | ARI | ARI | 0.92 | 1 |
| Basophils | Opt | ARI | 0.128 | 0.877 |
| Basophils | ARI | VHD | 0.92 | 1 |
| Basophils | Opt | VHD | 0.127 | 0.877 |
| Basophils | ARI | HD | 0.83 | 1 |
| Basophils | Opt | HD | 0.62 | 1 |
| Basophils | ARI | LD | 0.963 | 1 |
| Basophils | Opt | LD | 0.498 | 1 |
| Basophils | ARI | VLD | 0.181 | 0.931 |
| Basophils | Opt | VLD | 0.905 | 1 |
| Basophils | ARI | BRI | 0.83 | 1 |
| Basophils | Opt | BRI | 0.0654 | 0.727 |
| Bicarbonate | ARI | ARI | 0.384 | 1 |
| Bicarbonate | Opt | ARI | 0.623 | 1 |
| Bicarbonate | ARI | VHD | 0.588 | 1 |
| Bicarbonate | Opt | VHD | 4.22e-12 | 1.25e-09 |
| Bicarbonate | ARI | HD | 0.786 | 1 |
| Bicarbonate | Opt | HD | 0.588 | 1 |
| Bicarbonate | ARI | LD | 0.201 | 0.94 |
| Bicarbonate | Opt | LD | 0.82 | 1 |
| Bicarbonate | ARI | BRI | 0.182 | 0.931 |
| Bicarbonate | Opt | BRI | 0.723 | 1 |
| Bilirubin | ARI | ARI | 0.99 | 1 |
| Bilirubin | Opt | ARI | 0.327 | 1 |
| Bilirubin | ARI | VHD | 0.651 | 1 |
| Bilirubin | Opt | VHD | 0.184 | 0.931 |
| Bilirubin | ARI | HD | 0.667 | 1 |
| Bilirubin | Opt | HD | 0.457 | 1 |
| Bilirubin | ARI | LD | 0.682 | 1 |

| Parameter | CRP class | Drift class | p-value | BH-corrected p-value |
|---|---|---|---|---|
| Bilirubin | Opt | LD | 0.536 | 1 |
| Bilirubin | ARI | VLD | 0.469 | 1 |
| Bilirubin | Opt | VLD | 0.499 | 1 |
| Bilirubin | ARI | BRI | 0.1 | 0.788 |
| Bilirubin | Opt | BRI | 0.282 | 1 |
| Calcium | ARI | ARI | 0.877 | 1 |
| Calcium | Opt | ARI | 0.0456 | 0.612 |
| Calcium | ARI | HD | 0.182 | 0.931 |
| Calcium | Opt | HD | 0.417 | 1 |
| Calcium | ARI | LD | 0.562 | 1 |
| Calcium | Opt | LD | 0.36 | 1 |
| Calcium | ARI | VLD | 0.132 | 0.88 |
| Calcium | Opt | VLD | 0.165 | 0.931 |
| Calcium | ARI | BRI | 7.01e-04 | 0.0599 |
| Calcium | Opt | BRI | 0.0262 | 0.515 |
| Chloride | ARI | ARI | 0.264 | 1 |
| Chloride | Opt | ARI | 0.041 | 0.591 |
| Chloride | ARI | VHD | 0.397 | 1 |
| Chloride | Opt | VHD | 0.782 | 1 |
| Chloride | ARI | HD | 0.482 | 1 |
| Chloride | Opt | HD | 0.114 | 0.823 |
| Chloride | ARI | LD | 0.654 | 1 |
| Chloride | Opt | LD | 0.029 | 0.537 |
| Chloride | ARI | VLD | 0.461 | 1 |
| Chloride | Opt | VLD | 0.149 | 0.931 |
| Chloride | ARI | BRI | 0.161 | 0.931 |
| Chloride | Opt | BRI | 0.345 | 1 |
| Creatine kinase | ARI | ARI | 0.753 | 1 |
| Creatine kinase | Opt | ARI | 0.997 | 1 |
| Creatine kinase | ARI | VHD | 0.997 | 1 |
| Creatine kinase | Opt | VHD | 0.997 | 1 |
| Creatine kinase | ARI | HD | 0.997 | 1 |
| Creatine kinase | Opt | HD | 0.997 | 1 |
| Creatine kinase | ARI | LD | 0.376 | 1 |
| Creatine kinase | Opt | LD | 0.997 | 1 |
| Creatine kinase | ARI | VLD | 0.997 | 1 |
| Creatine kinase | Opt | VLD | 0.721 | 1 |
| Creatine kinase | ARI | BRI | 0.519 | 1 |
| Creatine kinase | Opt | BRI | 0.997 | 1 |
| Creatinine | ARI | ARI | 0.712 | 1 |
| Creatinine | Opt | ARI | 0.868 | 1 |
| Creatinine | ARI | VHD | 0.776 | 1 |
| Creatinine | Opt | VHD | 0.536 | 1 |
| Creatinine | ARI | HD | 0.384 | 1 |
| Creatinine | Opt | HD | 0.384 | 1 |
| Creatinine | ARI | LD | 0.279 | 1 |
| Creatinine | Opt | LD | 0.384 | 1 |
| Creatinine | ARI | VLD | 0.462 | 1 |
| Creatinine | Opt | VLD | 0.313 | 1 |
| Creatinine | ARI | BRI | 0.536 | 1 |
| Creatinine | Opt | BRI | 0.539 | 1 |
| ESR | ARI | ARI | 0.404 | 1 |
| ESR | Opt | ARI | 0.157 | 0.931 |
| ESR | ARI | VHD | 0.157 | 0.931 |
| ESR | ARI | HD | 0.796 | 1 |
| ESR | Opt | HD | 0.556 | 1 |
| ESR | ARI | BRI | 0.391 | 1 |
| ESR | Opt | BRI | 0.768 | 1 |
| Eosinophils | ARI | ARI | 0.771 | 1 |
| Eosinophils | Opt | ARI | 0.32 | 1 |
| Eosinophils | ARI | VHD | 0.619 | 1 |
| Eosinophils | Opt | VHD | 0.0903 | 0.783 |
| Eosinophils | ARI | HD | 0.923 | 1 |
| Eosinophils | Opt | HD | 0.0757 | 0.77 |
| Eosinophils | ARI | LD | 0.669 | 1 |
| Eosinophils | Opt | LD | 0.128 | 0.877 |
| Eosinophils | ARI | VLD | 0.419 | 1 |
| Eosinophils | Opt | VLD | 0.0124 | 0.351 |
| Eosinophils | ARI | BRI | 0.926 | 1 |
| Eosinophils | Opt | BRI | 0.181 | 0.931 |
| Fecal calprotectin | ARI | ARI | 0.0517 | 0.64 |
| Fecal calprotectin | Opt | ARI | 0.264 | 1 |
| Ferritin | ARI | ARI | 0.508 | 1 |
| Ferritin | Opt | ARI | 0.883 | 1 |
| Ferritin | ARI | VHD | 0.883 | 1 |
| Ferritin | Opt | VHD | 0.911 | 1 |
| Ferritin | ARI | HD | 0.985 | 1 |
| Ferritin | Opt | HD | 0.182 | 0.931 |
| Ferritin | ARI | LD | 0.533 | 1 |
| Ferritin | Opt | LD | 0.736 | 1 |
| Ferritin | ARI | BRI | 0.986 | 1 |

| Parameter | CRP class | Drift class | p-value | BH-corrected p-value |
|---|---|---|---|---|
| Ferritin | Opt | BRI | 0.95 | 1 |
| Fibrinogen | ARI | ARI | 0.239 | 1 |
| Fibrinogen | Opt | ARI | 0.339 | 1 |
| Fibrinogen | ARI | VHD | 0.997 | 1 |
| Fibrinogen | Opt | VHD | 0.919 | 1 |
| Fibrinogen | ARI | HD | 0.766 | 1 |
| Fibrinogen | Opt | HD | 0.773 | 1 |
| Fibrinogen | ARI | LD | 0.983 | 1 |
| Fibrinogen | Opt | LD | 0.209 | 0.953 |
| Fibrinogen | ARI | VLD | 0.855 | 1 |
| Fibrinogen | Opt | VLD | 0.796 | 1 |
| Fibrinogen | ARI | BRI | 8.93e-03 | 0.299 |
| Fibrinogen | Opt | BRI | 0.824 | 1 |
| Folate | ARI | VHD | 0.572 | 1 |
| Folate | Opt | VHD | 0.636 | 1 |
| Folate | ARI | HD | 0.792 | 1 |
| Folate | Opt | HD | 0.572 | 1 |
| Folate | ARI | BRI | 0.855 | 1 |
| Folate | Opt | BRI | 0.572 | 1 |
| Free T4 | ARI | ARI | 0.625 | 1 |
| Free T4 | Opt | ARI | 0.884 | 1 |
| Free T4 | ARI | VHD | 0.971 | 1 |
| Free T4 | Opt | VHD | 0.971 | 1 |
| Free T4 | ARI | HD | 0.468 | 1 |
| Free T4 | Opt | HD | 0.971 | 1 |
| Free T4 | ARI | LD | 0.52 | 1 |
| Free T4 | Opt | LD | 0.0981 | 0.788 |
| Free T4 | ARI | BRI | 0.169 | 0.931 |
| Free T4 | Opt | BRI | 6.34e-03 | 0.247 |
| GGT | ARI | ARI | 0.132 | 0.88 |
| GGT | Opt | ARI | 0.93 | 1 |
| GGT | ARI | VHD | 0.93 | 1 |
| GGT | Opt | VHD | 0.678 | 1 |
| GGT | ARI | HD | 0.968 | 1 |
| GGT | Opt | HD | 0.437 | 1 |
| GGT | ARI | LD | 0.678 | 1 |
| GGT | Opt | LD | 0.437 | 1 |
| GGT | ARI | VLD | 0.437 | 1 |
| GGT | Opt | VLD | 0.678 | 1 |
| GGT | ARI | BRI | 0.81 | 1 |
| GGT | Opt | BRI | 0.971 | 1 |
| Glucose | ARI | ARI | 0.925 | 1 |
| Glucose | Opt | ARI | 0.779 | 1 |
| Glucose | ARI | LD | 0.779 | 1 |
| Glucose | Opt | LD | 0.718 | 1 |
| Glucose | ARI | VLD | 0.943 | 1 |
| Glucose | Opt | VLD | 0.779 | 1 |
| Glucose | ARI | BRI | 0.779 | 1 |
| Glucose | Opt | BRI | 0.925 | 1 |
| Hematocrit | ARI | ARI | 0.663 | 1 |
| Hematocrit | Opt | ARI | 0.747 | 1 |
| Hematocrit | ARI | VHD | 0.629 | 1 |
| Hematocrit | Opt | VHD | 0.65 | 1 |
| Hematocrit | ARI | HD | 0.747 | 1 |
| Hematocrit | Opt | HD | 0.508 | 1 |
| Hematocrit | ARI | LD | 0.899 | 1 |
| Hematocrit | Opt | LD | 0.821 | 1 |
| Hematocrit | ARI | VLD | 0.493 | 1 |
| Hematocrit | Opt | VLD | 0.669 | 1 |
| Hematocrit | ARI | BRI | 0.0885 | 0.783 |
| Hematocrit | Opt | BRI | 0.207 | 0.953 |
| Hemoglobin | ARI | ARI | 0.903 | 1 |
| Hemoglobin | Opt | ARI | 0.201 | 0.94 |
| Hemoglobin | ARI | VHD | 0.749 | 1 |
| Hemoglobin | Opt | VHD | 0.993 | 1 |
| Hemoglobin | ARI | HD | 0.201 | 0.94 |
| Hemoglobin | Opt | HD | 0.0722 | 0.766 |
| Hemoglobin | ARI | LD | 0.62 | 1 |
| Hemoglobin | Opt | LD | 0.0941 | 0.785 |
| Hemoglobin | ARI | VLD | 0.201 | 0.94 |
| Hemoglobin | Opt | VLD | 0.978 | 1 |
| Hemoglobin | ARI | BRI | 0.0265 | 0.515 |
| Hemoglobin | Opt | BRI | 0.757 | 1 |
| Iron | ARI | ARI | 0.269 | 1 |
| Iron | Opt | ARI | 0.539 | 1 |
| Iron | ARI | VHD | 0.482 | 1 |
| Iron | Opt | VHD | 0.269 | 1 |
| Iron | ARI | HD | 0.31 | 1 |
| Iron | Opt | HD | 0.251 | 1 |
| Iron | ARI | LD | 0.355 | 1 |
| Iron | Opt | LD | 0.716 | 1 |

| Parameter | CRP class | Drift class | p-value | BH-corrected p-value |
|---|---|---|---|---|
| Iron | ARI | BRI | 0.0878 | 0.783 |
| Iron | Opt | BRI | 0.844 | 1 |
| LDH | ARI | ARI | 8.07e-04 | 0.0599 |
| LDH | Opt | ARI | 0.0238 | 0.515 |
| LDH | ARI | VHD | 0.275 | 1 |
| LDH | Opt | VHD | 0.922 | 1 |
| LDH | ARI | HD | 0.922 | 1 |
| LDH | Opt | HD | 0.613 | 1 |
| LDH | ARI | LD | 0.115 | 0.823 |
| LDH | Opt | LD | 0.855 | 1 |
| LDH | ARI | BRI | 0.0434 | 0.6 |
| LDH | Opt | BRI | 0.663 | 1 |
| LDL cholesterol | ARI | ARI | 0.273 | 1 |
| LDL cholesterol | Opt | ARI | 0.696 | 1 |
| LDL cholesterol | ARI | VHD | 0.54 | 1 |
| LDL cholesterol | Opt | VHD | 0.983 | 1 |
| LDL cholesterol | ARI | HD | 0.56 | 1 |
| LDL cholesterol | Opt | HD | 0.693 | 1 |
| LDL cholesterol | ARI | LD | 0.983 | 1 |
| LDL cholesterol | Opt | LD | 0.56 | 1 |
| LDL cholesterol | ARI | VLD | 0.56 | 1 |
| LDL cholesterol | Opt | VLD | 0.0612 | 0.713 |
| Lipase | ARI | ARI | 0.735 | 1 |
| Lipase | Opt | ARI | 0.324 | 1 |
| Lipase | ARI | HD | 0.552 | 1 |
| Lipase | Opt | HD | 1 | 1 |
| Lipase | ARI | LD | 0.552 | 1 |
| Lipase | Opt | LD | 0.552 | 1 |
| Lipase | ARI | VLD | 0.598 | 1 |
| Lipase | Opt | VLD | 0.574 | 1 |
| Lipase | ARI | BRI | 0.103 | 0.796 |
| Lipase | Opt | BRI | 0.574 | 1 |
| MCH | ARI | ARI | 0.895 | 1 |
| MCH | Opt | ARI | 0.271 | 1 |
| MCH | ARI | VHD | 0.314 | 1 |
| MCH | Opt | VHD | 0.702 | 1 |
| MCH | ARI | HD | 0.784 | 1 |
| MCH | Opt | HD | 0.271 | 1 |
| MCH | ARI | LD | 0.589 | 1 |
| MCH | Opt | LD | 0.169 | 0.931 |
| MCH | ARI | VLD | 0.115 | 0.823 |
| MCH | Opt | VLD | 0.79 | 1 |
| MCH | ARI | BRI | 0.482 | 1 |
| MCH | Opt | BRI | 0.271 | 1 |
| MCHC | ARI | ARI | 0.827 | 1 |
| MCHC | Opt | ARI | 0.732 | 1 |
| MCHC | ARI | VHD | 0.91 | 1 |
| MCHC | Opt | VHD | 0.99 | 1 |
| MCHC | ARI | HD | 0.732 | 1 |
| MCHC | Opt | HD | 0.99 | 1 |
| MCHC | ARI | LD | 0.916 | 1 |
| MCHC | Opt | LD | 0.0313 | 0.56 |
| MCHC | ARI | VLD | 0.968 | 1 |
| MCHC | Opt | VLD | 0.99 | 1 |
| MCHC | ARI | BRI | 0.239 | 1 |
| MCHC | Opt | BRI | 0.485 | 1 |
| MCV | ARI | ARI | 0.841 | 1 |
| MCV | Opt | ARI | 0.168 | 0.931 |
| MCV | ARI | VHD | 0.498 | 1 |
| MCV | Opt | VHD | 0.414 | 1 |
| MCV | ARI | HD | 0.675 | 1 |
| MCV | Opt | HD | 0.841 | 1 |
| MCV | ARI | LD | 0.701 | 1 |
| MCV | Opt | LD | 0.384 | 1 |
| MCV | ARI | VLD | 0.247 | 1 |
| MCV | Opt | VLD | 0.933 | 1 |
| MCV | ARI | BRI | 0.426 | 1 |
| MCV | Opt | BRI | 0.41 | 1 |
| Magnesium | ARI | ARI | 0.969 | 1 |
| Magnesium | Opt | ARI | 0.9 | 1 |
| Magnesium | ARI | VHD | 0.358 | 1 |
| Magnesium | Opt | VHD | 0.745 | 1 |
| Magnesium | ARI | HD | 0.969 | 1 |
| Magnesium | Opt | HD | 0.969 | 1 |
| Magnesium | ARI | LD | 0.803 | 1 |
| Magnesium | Opt | LD | 0.969 | 1 |
| Magnesium | ARI | VLD | 0.803 | 1 |
| Magnesium | Opt | VLD | 0.657 | 1 |
| Magnesium | ARI | BRI | 0.657 | 1 |
| Magnesium | Opt | BRI | 0.022 | 0.503 |
| Monocytes | ARI | ARI | 0.991 | 1 |

| Parameter | CRP class | Drift class | p-value | BH-corrected p-value |
|---|---|---|---|---|
| Monocytes | Opt | ARI | 0.271 | 1 |
| Monocytes | ARI | VHD | 0.995 | 1 |
| Monocytes | Opt | VHD | 0.252 | 1 |
| Monocytes | ARI | HD | 0.991 | 1 |
| Monocytes | Opt | HD | 0.0661 | 0.727 |
| Monocytes | ARI | LD | 0.644 | 1 |
| Monocytes | Opt | LD | 0.549 | 1 |
| Monocytes | ARI | VLD | 0.535 | 1 |
| Monocytes | Opt | VLD | 0.567 | 1 |
| Monocytes | ARI | BRI | 0.991 | 1 |
| Monocytes | Opt | BRI | 0.384 | 1 |
| Neutrophils | ARI | ARI | 0.0822 | 0.783 |
| Neutrophils | Opt | ARI | 0.997 | 1 |
| Neutrophils | ARI | VHD | 0.238 | 1 |
| Neutrophils | Opt | VHD | 0.959 | 1 |
| Neutrophils | ARI | HD | 0.357 | 1 |
| Neutrophils | Opt | HD | 0.946 | 1 |
| Neutrophils | ARI | LD | 0.35 | 1 |
| Neutrophils | Opt | LD | 0.196 | 0.94 |
| Neutrophils | ARI | VLD | 0.56 | 1 |
| Neutrophils | Opt | VLD | 0.0412 | 0.591 |
| Neutrophils | ARI | BRI | 0.997 | 1 |
| Neutrophils | Opt | BRI | 0.253 | 1 |
| Phosphate | ARI | ARI | 0.996 | 1 |
| Phosphate | Opt | ARI | 0.477 | 1 |
| Phosphate | ARI | VHD | 0.917 | 1 |
| Phosphate | Opt | VHD | 0.555 | 1 |
| Phosphate | ARI | HD | 0.668 | 1 |
| Phosphate | Opt | HD | 0.377 | 1 |
| Phosphate | ARI | LD | 0.206 | 0.953 |
| Phosphate | Opt | LD | 0.189 | 0.935 |
| Phosphate | ARI | VLD | 0.477 | 1 |
| Phosphate | Opt | VLD | 0.919 | 1 |
| Phosphate | ARI | BRI | 0.153 | 0.931 |
| Phosphate | Opt | BRI | 0.917 | 1 |
| Platelets | ARI | ARI | 4.20e-04 | 0.0499 |
| Platelets | Opt | ARI | 0.449 | 1 |
| Platelets | ARI | VHD | 0.0369 | 0.588 |
| Platelets | Opt | VHD | 0.926 | 1 |
| Platelets | ARI | HD | 0.343 | 1 |
| Platelets | Opt | HD | 0.944 | 1 |
| Platelets | ARI | LD | 0.706 | 1 |
| Platelets | Opt | LD | 0.694 | 1 |
| Platelets | ARI | BRI | 0.855 | 1 |
| Platelets | Opt | BRI | 0.85 | 1 |
| Potassium | ARI | ARI | 0.6 | 1 |
| Potassium | Opt | ARI | 0.224 | 1 |
| Potassium | ARI | VHD | 0.647 | 1 |
| Potassium | Opt | VHD | 0.174 | 0.931 |
| Potassium | ARI | HD | 0.185 | 0.931 |
| Potassium | Opt | HD | 0.0905 | 0.783 |
| Potassium | ARI | LD | 0.6 | 1 |
| Potassium | Opt | LD | 0.147 | 0.931 |
| Potassium | ARI | VLD | 0.634 | 1 |
| Potassium | Opt | VLD | 0.519 | 1 |
| Potassium | ARI | BRI | 4.03e-03 | 0.218 |
| Potassium | Opt | BRI | 0.162 | 0.931 |
| Red blood cells | ARI | ARI | 0.451 | 1 |
| Red blood cells | Opt | ARI | 0.94 | 1 |
| Red blood cells | ARI | VHD | 0.96 | 1 |
| Red blood cells | Opt | VHD | 0.316 | 1 |
| Red blood cells | ARI | HD | 0.399 | 1 |
| Red blood cells | Opt | HD | 0.394 | 1 |
| Red blood cells | ARI | LD | 0.693 | 1 |
| Red blood cells | Opt | LD | 0.751 | 1 |
| Red blood cells | ARI | VLD | 0.32 | 1 |
| Red blood cells | Opt | VLD | 0.35 | 1 |
| Red blood cells | ARI | BRI | 0.311 | 1 |
| Red blood cells | Opt | BRI | 0.519 | 1 |
| Reticulocytes | ARI | ARI | 0.826 | 1 |
| Reticulocytes | Opt | ARI | 0.323 | 1 |
| Reticulocytes | ARI | VHD | 0.483 | 1 |
| Reticulocytes | Opt | VHD | 0.508 | 1 |
| Reticulocytes | ARI | HD | 0.968 | 1 |
| Reticulocytes | Opt | HD | 0.724 | 1 |
| Reticulocytes | ARI | LD | 0.648 | 1 |
| Reticulocytes | Opt | LD | 0.958 | 1 |
| Reticulocytes | ARI | VLD | 0.999 | 1 |
| Reticulocytes | Opt | VLD | 2.17e-14 | 1.29e-11 |
| Reticulocytes | ARI | BRI | 0.92 | 1 |
| Reticulocytes | Opt | BRI | 0.999 | 1 |

| Parameter | CRP class | Drift class | p-value | BH-corrected p-value |
|---|---|---|---|---|
| Sodium | ARI | ARI | 0.0649 | 0.727 |
| Sodium | Opt | ARI | 0.0465 | 0.612 |
| Sodium | ARI | VHD | 0.978 | 1 |
| Sodium | Opt | VHD | 0.844 | 1 |
| Sodium | ARI | HD | 0.377 | 1 |
| Sodium | Opt | HD | 0.7 | 1 |
| Sodium | ARI | LD | 0.647 | 1 |
| Sodium | Opt | LD | 7.62e-04 | 0.0599 |
| Sodium | ARI | VLD | 0.978 | 1 |
| Sodium | Opt | VLD | 3.81e-03 | 0.218 |
| Sodium | ARI | BRI | 0.498 | 1 |
| Sodium | Opt | BRI | 0.244 | 1 |
| TSH | ARI | ARI | 0.874 | 1 |
| TSH | Opt | ARI | 0.963 | 1 |
| TSH | ARI | VHD | 0.748 | 1 |
| TSH | Opt | VHD | 0.963 | 1 |
| TSH | ARI | HD | 0.991 | 1 |
| TSH | Opt | HD | 0.963 | 1 |
| TSH | ARI | LD | 0.874 | 1 |
| TSH | Opt | LD | 0.986 | 1 |
| TSH | ARI | VLD | 0.963 | 1 |
| TSH | Opt | VLD | 0.941 | 1 |
| TSH | ARI | BRI | 0.991 | 1 |
| TSH | Opt | BRI | 0.748 | 1 |
| Total cholesterol | ARI | ARI | 0.312 | 1 |
| Total cholesterol | Opt | ARI | 0.463 | 1 |
| Total cholesterol | ARI | HD | 0.101 | 0.788 |
| Total cholesterol | Opt | HD | 0.321 | 1 |
| Total cholesterol | ARI | LD | 0.164 | 0.931 |
| Total cholesterol | Opt | LD | 0.0151 | 0.389 |
| Total cholesterol | ARI | VLD | 0.0956 | 0.785 |
| Total cholesterol | Opt | VLD | 5.50e-03 | 0.247 |
| Total cholesterol | ARI | BRI | 0.0108 | 0.321 |
| Total cholesterol | Opt | BRI | 0.0343 | 0.575 |
| Total lymphocytes | ARI | ARI | 0.163 | 0.931 |
| Total lymphocytes | Opt | ARI | 0.551 | 1 |
| Total lymphocytes | ARI | VHD | 0.278 | 1 |
| Total lymphocytes | Opt | VHD | 0.166 | 0.931 |
| Total lymphocytes | ARI | HD | 0.234 | 1 |
| Total lymphocytes | Opt | HD | 0.163 | 0.931 |
| Total lymphocytes | ARI | LD | 0.792 | 1 |
| Total lymphocytes | Opt | LD | 0.693 | 1 |
| Total lymphocytes | ARI | VLD | 0.596 | 1 |
| Total lymphocytes | Opt | VLD | 0.849 | 1 |
| Total lymphocytes | ARI | BRI | 0.64 | 1 |
| Total lymphocytes | Opt | BRI | 0.353 | 1 |
| Total proteins | ARI | ARI | 0.661 | 1 |
| Total proteins | Opt | ARI | 0.661 | 1 |
| Total proteins | ARI | VHD | 0.905 | 1 |
| Total proteins | Opt | VHD | 0.0949 | 0.785 |
| Total proteins | ARI | HD | 0.661 | 1 |
| Total proteins | Opt | HD | 0.0879 | 0.783 |
| Total proteins | ARI | LD | 0.0481 | 0.612 |
| Total proteins | Opt | LD | 0.935 | 1 |
| Total proteins | ARI | VLD | 0.765 | 1 |
| Total proteins | Opt | VLD | 0.905 | 1 |
| Total proteins | ARI | BRI | 0.016 | 0.396 |
| Total proteins | Opt | BRI | 0.0149 | 0.389 |
| Transferrin | ARI | ARI | 0.318 | 1 |
| Transferrin | Opt | ARI | 0.933 | 1 |
| Transferrin | ARI | VHD | 0.971 | 1 |
| Transferrin | Opt | VHD | 0.728 | 1 |
| Transferrin | ARI | HD | 0.909 | 1 |
| Transferrin | Opt | HD | 0.776 | 1 |
| Transferrin | ARI | LD | 0.707 | 1 |
| Transferrin | Opt | LD | 0.994 | 1 |
| Transferrin | ARI | BRI | 0.0909 | 0.783 |
| Transferrin | Opt | BRI | 0.909 | 1 |
| Triglycerides | ARI | ARI | 0.788 | 1 |
| Triglycerides | Opt | ARI | 0.622 | 1 |
| Triglycerides | ARI | VHD | 0.622 | 1 |
| Triglycerides | Opt | VHD | 0.838 | 1 |
| Triglycerides | ARI | HD | 0.647 | 1 |
| Triglycerides | Opt | HD | 0.583 | 1 |
| Triglycerides | ARI | LD | 0.501 | 1 |
| Triglycerides | Opt | LD | 0.622 | 1 |
| Triglycerides | ARI | VLD | 0.622 | 1 |
| Urea | ARI | ARI | 0.356 | 1 |
| Urea | Opt | ARI | 0.732 | 1 |
| Urea | ARI | VHD | 0.849 | 1 |
| Urea | Opt | VHD | 0.328 | 1 |

| Parameter | CRP class | Drift class | p-value | BH-corrected p-value |
|---|---|---|---|---|
| Urea | ARI | HD | 0.548 | 1 |
| Urea | Opt | HD | 0.179 | 0.931 |
| Urea | ARI | LD | 0.816 | 1 |
| Urea | Opt | LD | 0.431 | 1 |
| Urea | ARI | BRI | 0.419 | 1 |
| Urea | Opt | BRI | 0.752 | 1 |
| Uric acid | ARI | ARI | 0.249 | 1 |
| Uric acid | Opt | ARI | 0.636 | 1 |
| Uric acid | ARI | VHD | 0.709 | 1 |
| Uric acid | Opt | VHD | 0.22 | 0.989 |
| Uric acid | ARI | HD | 0.235 | 1 |
| Uric acid | Opt | HD | 0.187 | 0.931 |
| Uric acid | ARI | LD | 0.437 | 1 |
| Uric acid | Opt | LD | 0.479 | 1 |
| Uric acid | ARI | BRI | 0.241 | 1 |
| Uric acid | Opt | BRI | 0.433 | 1 |
| Vitamin B12 | ARI | ARI | 0.982 | 1 |
| Vitamin B12 | Opt | ARI | 0.568 | 1 |
| Vitamin B12 | ARI | HD | 0.468 | 1 |
| Vitamin B12 | Opt | HD | 0.952 | 1 |
| Vitamin B12 | ARI | LD | 0.944 | 1 |
| Vitamin B12 | Opt | LD | 0.468 | 1 |
| Vitamin B12 | ARI | VLD | 0.568 | 1 |
| Vitamin B12 | Opt | VLD | 0.944 | 1 |
| Vitamin B12 | ARI | BRI | 0.944 | 1 |
| Vitamin B12 | Opt | BRI | 0.468 | 1 |
| Vitamin D | ARI | ARI | 0.418 | 1 |
| Vitamin D | Opt | ARI | 0.756 | 1 |
| Vitamin D | ARI | HD | 0.395 | 1 |
| Vitamin D | Opt | HD | 0.395 | 1 |
| Vitamin D | ARI | BRI | 0.395 | 1 |
| Vitamin D | Opt | BRI | 0.232 | 1 |
| White blood cells | ARI | ARI | 0.342 | 1 |
| White blood cells | Opt | ARI | 0.858 | 1 |
| White blood cells | ARI | VHD | 0.573 | 1 |
| White blood cells | Opt | VHD | 0.0379 | 0.588 |
| White blood cells | ARI | HD | 0.702 | 1 |
| White blood cells | Opt | HD | 0.763 | 1 |
| White blood cells | ARI | LD | 0.683 | 1 |
| White blood cells | Opt | LD | 0.0964 | 0.785 |
| White blood cells | ARI | VLD | 0.769 | 1 |
| White blood cells | Opt | VLD | 0.616 | 1 |
| White blood cells | ARI | BRI | 0.831 | 1 |
| White blood cells | Opt | BRI | 0.623 | 1 |
